# An integrated framework for functional dissection of variable expressivity in genetic disorders

**DOI:** 10.1101/2025.07.22.25331885

**Authors:** Jiawan Sun, Serena Noss, Deepro Banerjee, Venkata Hemanjani Bhavana, Corrine Smolen, Maitreya Das, Belinda Giardine, Anisha Prabhu, David J Amor, Kate Pope, Paul J Lockhart, Santhosh Girirajan

## Abstract

Disease-associated variants can lead to variable phenotypic outcomes, but the biological mechanisms underlying this variability remain poorly understood. We developed a framework to investigate this phenomenon using the 16p12.1 deletion as a paradigm of variable expressivity. Using induced pluripotent stem cell models from affected families and CRISPR-edited lines with the 16p12.1 deletion, we found that the deletion and secondary variants in the genetic background jointly influenced chromatin accessibility and expression of neurodevelopmental genes. Cellular analyses identified family-specific phenotypes, including altered inhibitory neuron production and neural progenitor cell proliferation, which correlated with head-size variation. CRISPR activation of individual 16p12.1 genes variably rescued these defects by modulating key signaling pathways such as TGF-β and PI3K-AKT. Integrative analyses further identified regulatory hubs, including transcription factors FOXG1 and JUN, as mediators of these effects. Our study provides a functional framework for investigating how individual genetic architectures contribute to phenotypic variability in genetic disorders.

## INTRODUCTION

In contrast to Mendelian disorders with straightforward genotype to phenotype relationships, variants associated with complex disorders often lead to variable clinical features^1, 2^. This phenomenon of variable expressivity is particularly true for rare copy number variants (CNVs), such as deletions and duplications within chromosomal regions 1q21.1, 15q13.3, and 22q11.2, which have been associated with variable neurodevelopmental outcomes and have also been identified in population controls^3^. For example, the 22q11.2 deletion, classically known to cause DiGeorge syndrome, is associated with intellectual disability and developmental delay (ID/DD), autism, and congenital heart defects, and it accounts for about 1% of individuals with schizophrenia^4, 5^. The approximately 500-kbp 16p12.1 deletion, encompassing eight genes, represents an ideal model to study variable expressivity of disease-associated variants for several reasons. *First*, the deletion is associated with a wide range of phenotypic outcomes, including ID/DD, autism, congenital anomalies, and epilepsy in affected children, as well as psychiatric features such as schizophrenia, depression, and anxiety in adolescents and adults, with varying levels of severity^6–12^. *Second*, unlike syndromic CNVs such as 17p11.2 deletion in Smith-Magenis syndrome or 5q35 deletion in Sotos syndrome^3^, that mostly occur *de novo*, the 16p12.1 deletion is inherited in over 90% of affected individuals from parents with milder cognitive or neuropsychiatric features^13^. The high inheritance rate allows us to assess phenotypic outcomes in multiple carriers from the same family. *Third*, specific patterns of rare variants in the genetic background (“secondary variants”) correlated with distinct phenotypic trajectories associated with the deletion^14^. In fact, modifier roles of secondary variants have also been reported as contributing to phenotypic variability in both monogenic and complex neurodevelopmental disorders, including in individuals affected by 7q11.23 duplication and 16p11.2 duplication ^2, 3, 15, 16^. However, the molecular basis by which these CNVs confer disease susceptibility and interact with secondary variants to produce variable outcomes remains poorly understood.

Previous studies have shown the utility of induced pluripotent stem cells (iPSC) to recapitulate complex biological processes *in vitro*, enabling investigations into the molecular etiology of neurodevelopmental disorders^17–19^. However, these studies have typically focused on aggregate analysis of subjects with the same primary variant, such as the 16p11.2 and 22q11.2 deletion, or the same phenotype, such as autism and schizophrenia, without investigating the variability among these subjects^20–22^. Thus, the effects of disease-associated variants within the context of an individual’s genetic background remain unexplored. Here, we developed a framework to dissect the mechanistic basis of variable expressivity in iPSC models of the 16p12.1 deletion **(Figure 1)**. By integrating multi-omics profiling, CRISPR/dCas9-mediated gene activation, and family-based analyses, we demonstrate that phenotypic trajectories associated with 16p12.1 deletion are shaped by the combinatorial effects of the deletion and secondary variants through the modulation of key signaling pathways. We further identified regulatory hubs that mediate these effects, including transcription factors (TFs) FOXG1 and JUN, along with genes in the regulatory network. We propose a conceptual shift from a focus on single causal variants to a system-level understanding of individual genetic architecture, advancing more effective precision medicine strategies.

**Figure 1.**
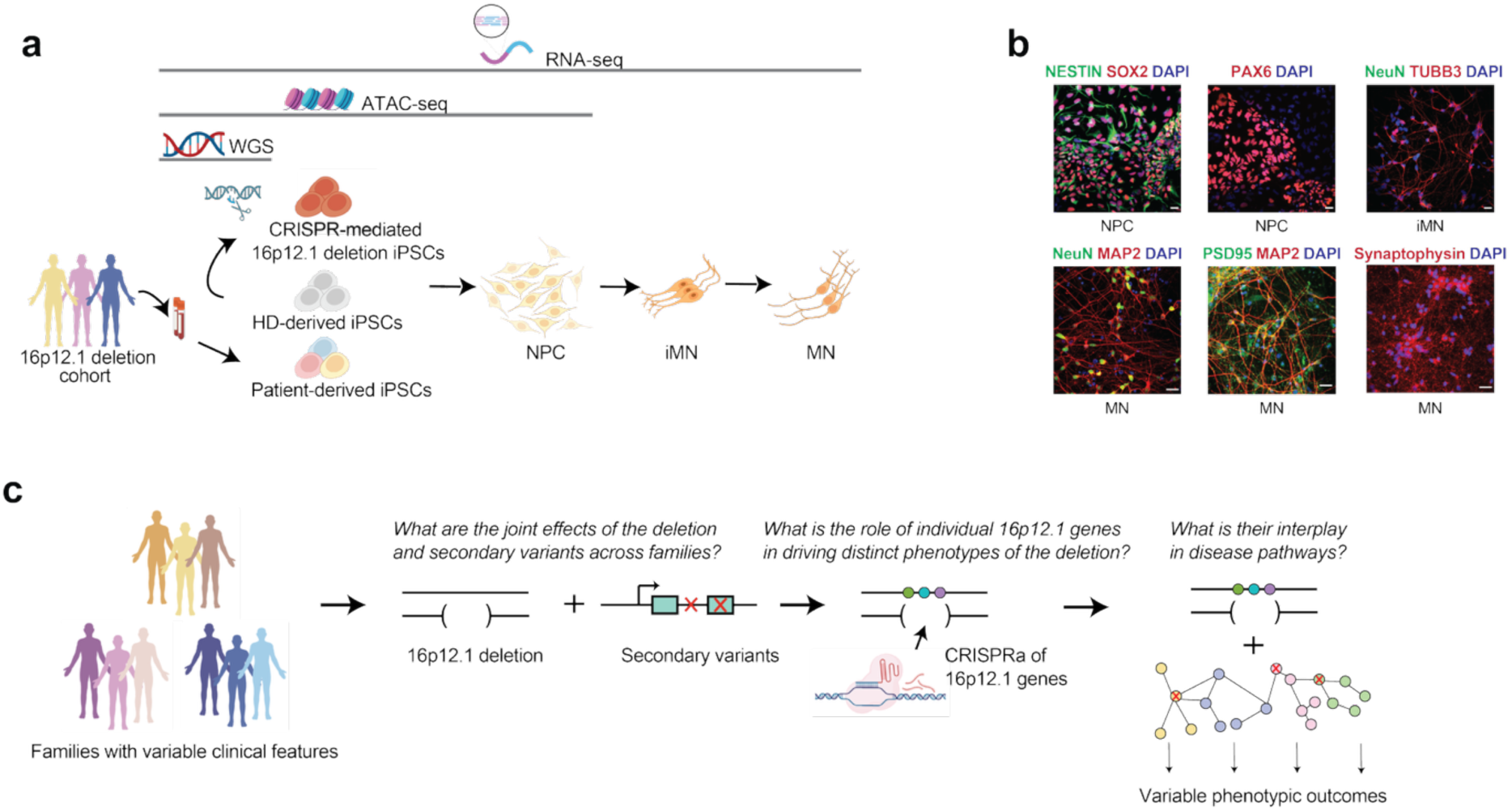
Overview of the integrated framework using 16p12.1 deletion iPSC models. (**a**) Experimental design. WGS was performed on DNA isolated from blood or iPSCs derived from healthy donors and individuals in the three families. RNA-seq was performed across all four differentiation stages and ATAC-seq was performed at iPSC and NPC stages. (**b**) Representative images of neural-converted cells stained with different markers for distinct cell stages (scale bars, 20 μm). **(c)** Overview of experimental strategy used to investigate how the 16p12.1 deletion and secondary variants contribute to phenotypic variability. The core questions driving the experiments within the framework are also shown.

## RESULTS

### Generation and characterization of iPSC models for 16p12.1 deletion

To model the effects of the deletion on neuronal development in an isogenic setting, we created a 465-kbp 16p12.1 deletion in the HD_01 line, derived from a healthy donor, using a CRISPR/Cas-9-based strategy **(Supplementary Figure 1A-E)**. To study the effects of the 16p12.1 deletion under different family-specific genetic backgrounds, we reprogrammed iPSC lines from peripheral blood mononuclear cells derived from 12 individuals from three families **(Figure 1A-B).** As controls, we used iPSC lines (HD_01 and HD_02; obtained from NINDS repository) from unrelated healthy male and female donors, as well as a HD_01 line transfected with empty vector as a CRISPR control. These 16 iPSC lines were then differentiated to neural progenitor cells (NPC) and immature (iMN) and mature neurons (MN) using dual-SMAD inhibition^18^ (see **Methods, Figure 1B)**. RNA sequencing was performed at all four cell stages, while ATAC-seq was performed for iPSC and NPC lines (**Figure 1A**). We confirmed reduced expression of 16p12.1 deletion genes including *UQCRC2, POLR3E, MOSMO*, and *CDR2*, as well as reduced ATAC-seq signals across the region **(Supplementary Figure 2A-B, Supplementary Table 1-2).** The expression of expected gene markers were robust for each differentiation cell state, such as expression of *SOX2*, *POU5F1*, and *FUT4* for iPSC; *SOX2*, *HES1*, *NES*, and *EMX2* for NPC; and *MAP2*, *DLG4*, *NCAM1*, and *RBFOX3* for neurons **(Supplementary Figure 2C)**, and ATAC-seq peak revealed differences in chromatin accessibility near cell-type-specific genetic markers such as *POU5F1*, *SOX2,* and *HES1*, confirming the expected cell identities across the lines **(Supplementary Figure 2D).**

### Combined effects on gene expression related to neurodevelopment

We compared all deletion lines to nondeletion lines at each cell stage and found that the most significantly differentially expressed genes (DEGs) were those within the deleted region (**Supplementary Figure 2A, Supplementary Table 1)**. This suggested that a combined analysis may obscure the downstream effects of the deletion due to variability between samples, likely due to differences in their genetic background. To isolate the direct impact of the deletion, we compared the CRISPR-edited 16p12.1 deletion line to its isogenic control. This approach is commonly used in iPSC models of disease^23^ to control for the effects of the genetic background. DEGs identified in NPC, iMN, and MN were enriched for neurodevelopmental and psychiatric disorder genes from the DisGeNET and published datasets^24–29^ **(Supplementary Figure 1F-G)**. To assess the extent to which the effects of genetic background on DEGs were controlled in the isogenic setting, we performed whole genome sequencing and identified 6,659 rare variants (referred to as *secondary variants*) in the HD_01 line (see **Methods**). We classified these secondary variants into distinct classes, including CNVs, simple tandem repeat (STR) expansions, and SNVs within exonic, intronic, upstream, downstream, 5’, and 3’ UTR regions of genes, as previously described^14, 30^, and found that 2%, 31%, 21%, and 10% of secondary variants overlapped DEGs identified in iPSC, NPC, iMN, and MN, respectively **(Figure 2A, Supplementary Table 1)**. Rare noncoding intronic variants were also significantly associated with altered usage of isoforms in genes without expression changes (**Figure 2A, Supplementary Table 1)**. Over-representation analysis of genes and isoforms with secondary variants revealed significant enrichment for neurodevelopment, neuronal function, and signaling pathways such as axon guidance, synaptic signaling, and PI3K-AKT signaling pathway (**Supplementary Figure 1H, Supplementary Table 1)**. These results suggest that secondary variants, though individually rare but collectively prevalent in healthy populations^31^, can alter gene expression when combined with the 16p12.1 deletion^32^. This observation also supports the idea that secondary variants may exert latent effects that become unmasked in the context of the deletion^33, 34^.

**Figure 2.**
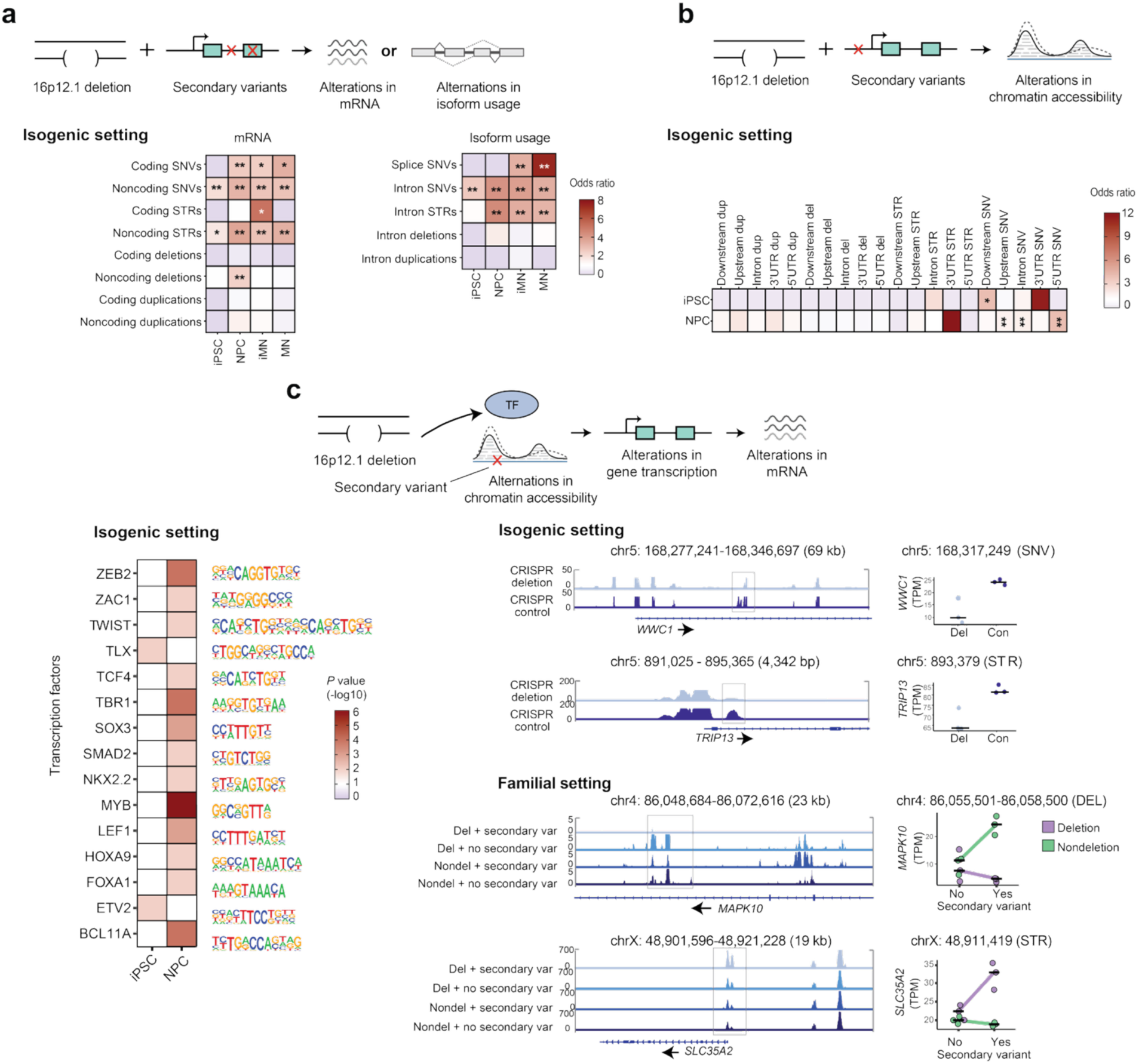
Evaluating the impact of secondary variants on chromatin accessibility and gene expression in the context of 16p12.1 deletion. (**a**) Overlap between secondary variants and DEGs (padj<0.05) or isoforms with altered usages (q<0.05) in the isogenic setting. Color represents odds ratio from Fisher’s exact test. *p<0.05, **Benjamini-Hochberg FDR<0.05, Fisher’s exact test. (**b**) Overlap between secondary variants and differential peaks obtained from ATAC-seq (padj<0.01) in the isogenic setting. Color represents odds ratio from Fisher’s exact test. *p<0.05, **Benjamini-Hochberg FDR<0.05, Fisher’s exact test. **(c)** Representative motifs (left) of transcription factor binding sites enriched within ATAC-seq differential peak regions that intersect with secondary variants. Examples (right) of altered chromatin accessibility peaks that intersect with secondary variants as well as associated differential expression of the corresponding gene in the isogenic and familial settings (transcripts per million, TPM, with median values) at the NPC stages. ATAC-seq peaks were visualized using Integrative Genome Viewer (IGV). From the DESeq2 results, the adjusted p-value (padj) for *WWC1* was 0.00041, and for *TRIP13*, padj was 0.018 **(Supplementary Table 8)**. Con, CRISPR control; Del, CRISPR deletion; var, variant; DEL, deletion; SNV, single nucleotide variant; STR, short tandem repeat; Del, 16p12.1 deletion lines; Nondel, nondeletion lines. Data in the familial setting were derived from the GL_079 samples **(Supplementary Table 3).** For both RNA-seq and ATAC-seq, we used n=3 independent experiments.

Given the significant overlap between rare noncoding variants and DEGs (**Figure 2A**), we evaluated their potential to alter chromatin accessibility. We identified six and 130 rare noncoding variants that intersected with differentially accessible chromatin (i.e., differential peaks or “Diffpeak”) regions from ATAC-seq analysis of iPSCs and NPCs, respectively **(Figure 2B, Supplementary Table 2).** Motif enrichment analysis of these intersecting Diffpeak regions revealed significant enrichment for binding motifs of TFs such as FOXA1, MYB, and TCF4, which are involved in signaling pathways such as TGF-β, Wnt, and PI3K-AKT that are relevant to neurodevelopment^35–38^ (**Figure 2C, Supplementary Table 2)**. We also observed differential expression of target genes affected by these noncoding secondary variants, such as *WWC1* and *TRIP13*, which are likely involved in a broader regulatory network of those TFs (**Figure 2C**).

We next tested the combined effects of the 16p12.1 deletion and the secondary variants using iPSC models derived from families with variable clinical features **(Figure 1C).** The DEGs identified in deletion lines compared to nondeletion lines from these families were enriched for genes associated with neurodevelopmental and psychiatric disorders; however, the enrichment patterns varied by family (**Supplementary Figure 3A-B, Supplementary Table 8)**. We analyzed each family separately and found that genes with secondary variants, including *MAPK10* and *SLC35A2*, showed significant changes in chromatin accessibility and gene expression when both the deletion and secondary variant were present, suggesting potential nonadditive effects. (**Figure 2C, Supplementary Figure 3C Supplementary Table 3).** This phenomenon has been reported previously, where nonadditive functional effects were observed among multiple schizophrenia-associated common variants in iPSC models^39, 40^. Overall, our findings support a modifier role for secondary variants, which act in combination with the deletion, to shape gene expression patterns within neurodevelopmental pathways.

### Variable neuronal phenotypes across genetic backgrounds

Beyond transcriptomic changes, we also evaluated the combined effects of the deletion and secondary variants on neuronal phenotypes. At the NPC stage, we found no differences in NESTIN and SOX2 levels between the deletion and nondeletion lines. However, we observed a decrease in PAX6 (a dorsal telencephalic marker) (**Supplementary Figure 4 A-B)** and a highly variable increase in NKX2.1 (a ventral telencephalic marker) in the deletion lines compared to nondeletion lines **(Figure 3A),** suggesting altered neuronal lineage commitment during differentiation. To further investigate this variability, we performed family-specific analysis comparing the deletion lines to HD lines and, when available, to nondeletion lines from the same family. We found a marked increase in NKX2.1-positive cells specifically in GL_007 and the CRISPR deletion lines **(Figure 3A-B)**. These results are consistent with previous findings in other iPSC models of neurodevelopmental disorders^41^. In addition to NKX2.1, which is critical for the development of inhibitory neurons, we examined VGAT, the vesicular GABA transporter, in mature neurons expressing NeuN and MAP2 (**Supplementary Figure 4C)** and found opposing trends across families, with a substantial increase in VGAT intensity in the deletion lines of GL_007 and a variable decrease in the deletion lines of GL_077 and GL_079 compared to controls **(Figure 3C-D)**. We also assessed other genes involved in inhibitory neuron differentiation and migration, including *DLX1*, *DLX2*, *LHX6* and *LHX8*, and found them to be highly expressed across NPCs, iMNs, and MNs in both the CRISPR deletion and GL_007 deletion lines compared to controls (**Supplementary Figure 4D, Supplementary Tables 8)**. Although NKX2.1 level was elevated in the CRISPR deletion line compared to its isogenic control, the absence of a significant difference in VGAT intensity suggests that this alteration may not persist into the MN stage or may be too subtle to be detected with this assay.

**Figure 3.**
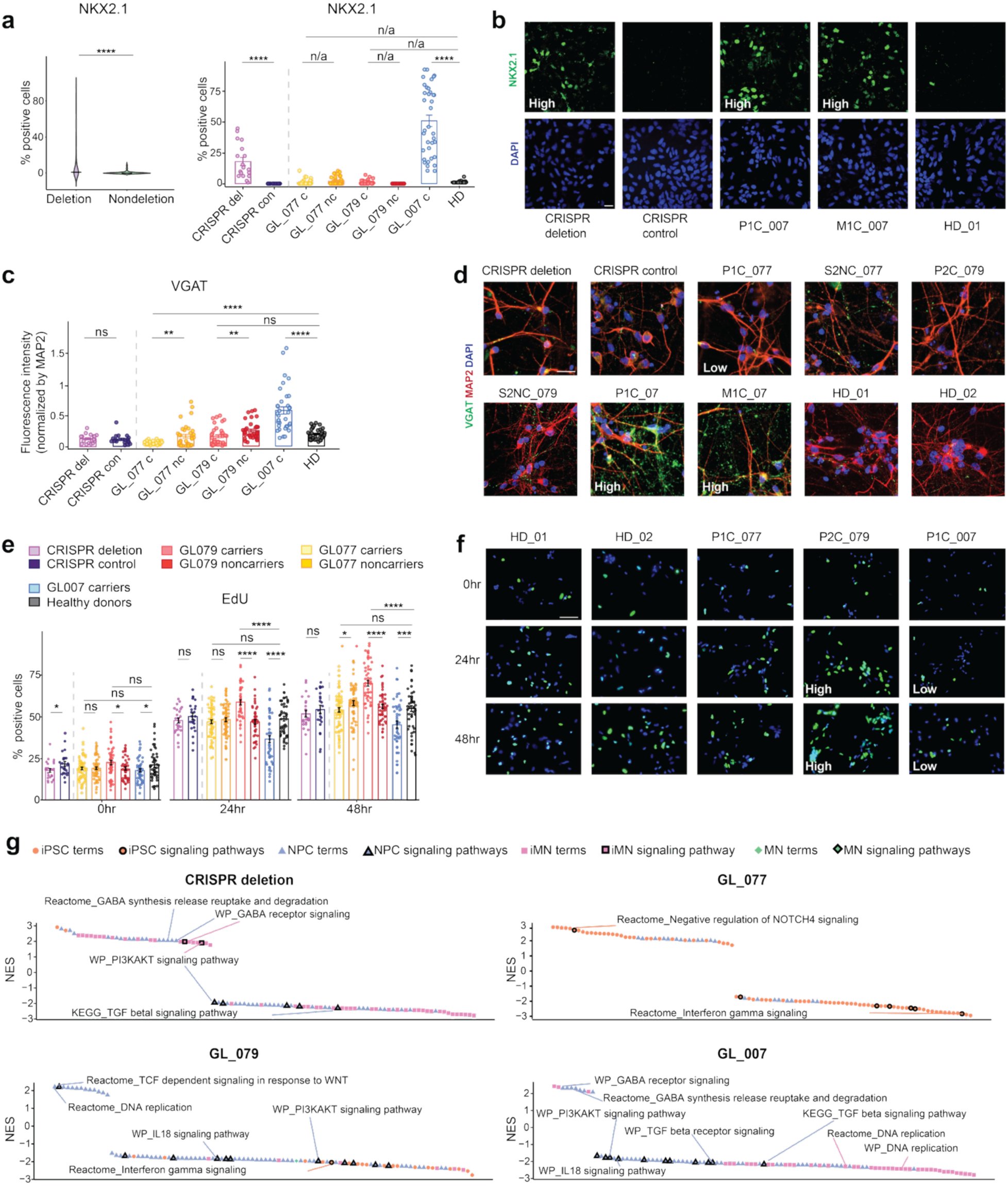
Assessing neuronal phenotypes across genetic backgrounds. **(a)** Violin plot (left) with median values shows the percentage of NKX2.1-positive cells at the NPC stage in all deletion lines compared to all nondeletion lines. Bar plot (right) with mean values shows the percentage of NKX2.1-positive cells within family-specific lines. **(b)** Representative images of NPCs stained for NKX2.1. Scale bars, 20 μm. **(c)** Bar plot with mean values shows VGAT fluorescence intensity normalized by MAP2 fluorescence intensity in each image. **(d)** Representative images of neurons stained for VGAT and MAP2. Scale bars, 20 μm. **(e)** Bar plot with mean values shows the percentage of Edu-positive cells in specific groups. **(f)** Representative images of EdU-labeled NPCs across three timepoints. Scale bars, 70 μm. **(g)** Waterfall plot of unbiasedly selected top pathways from each comparison of deletion versus nondeletion samples using GSEA (q<0.05) annotated in the Reactome, KEGG and WikiPathway (WP) (see **Methods**). NES, normalized enrichment score; isogenic, CRISPR deletion versus CRISPR control. Comparisons involve the grouped deletion carriers of each family versus the two healthy donor lines. The bold border indicates terms that belong to signaling pathways. Data are mean±s.e.m and two-tailed Welch’s t-tests for **(a), (c), (e)**. *p<0.05; **p<0.01; ***p<0.001; ****p<0.0001. n=3 independent experiments for each line, except for RNA-seq and downstream analyses where n=2 for iMNs and MNs of P1C_077, and MNs of HD_01. “n/a” indicates that Welch’s t-tests were not performed due to high zero inflation. For **(a)** and **(c)**, n=6 images per experiment and for **(e)** n=8 images per experiment were quantified. For **(a)** and **(e)**, for GL_079, GL_077, GL_007, “c” contains all deletion lines and “nc” contains all nondeletion lines, for their respective families (see Figure 1C). For **(c)**, the same cell lines were used, excluding GMC_077 and GFNC_077 lines. In the family-based analyses, we compared deletion lines from GL_077 and GL_079 to both nondeletion lines within the same family and HD lines. For GL_007, due to the absence of nondeletion family lines, we compared the deletion lines to HD lines.

Additionally, glutamatergic neuron production did not appear to be affected, as expression levels of glutamatergic markers (*SLC17A6*, *SLC17A7*, *GRIN1*, *GRIN2B*, and *GLS*) were not consistently altered across the deletion lines, and mature neurons showed no overt change in VGLUT1 expression **(Supplementary Figure 4C)**. These results suggest that the deletion contributes to altered production of inhibitory neurons under specific genetic backgrounds. Aberrant neuronal proliferation and apoptosis, which alter the proportions of distinct brain cell types, represent another convergent phenotype in neurodevelopmental disorders^42^. We evaluated NPC proliferation by measuring EdU-labeled cells at 0, 24, and 48 hours, as well as Ki-67-positive cells, and found no differences in proliferation rates between the deletion and nondeletion lines (**Supplementary Figure 5A-B)**. Similarly, TUNEL assays revealed no differences in apoptosis between the two groups (**Supplementary Figure 5C**). However, when grouped by family, we observed opposite trends in the deletion lines from GL_079 and GL_007 compared to controls: GL_079 deletion lines showed increased proliferation, while GL_007 deletion lines showed decreased proliferation and increased apoptosis compared to HD controls (**Figure 3E-F, Supplementary Figure 5D-G)**. GL_079 deletion lines also showed decreased apoptosis, but only when compared to nondeletion lines from the same family **(Supplementary Figure 5F)**. Notably, these results correspond with the head-size phenotypes in P2C_079 and FNC_079 from family GL_079 (macrocephaly) and P1C_007 from family GL_007 (microcephaly).

We also found evidence of altered developmental timing in deletion lines, a phenomenon also reported in iPSC models of autism and schizophrenia^43, 44^. A significant increase in TUBB3-positive cells (a marker for newborn neurons) at the NPC stage was detected in the CRISPR deletion and GL_007 deletion lines compared to controls **(Supplementary Figure 5H-J).** To further investigate this trend, we performed weighted gene co-expression network analysis (WGCNA) on RNA-seq data from NPC, iMN, and MN stages to identify gene modules correlated with specific stages of differentiation and examine changes in these modules within each family (**Supplementary Figure 6A, Supplementary Table 7**). These gene modules were enriched for differentiation stage-specific Gene Ontology (GO) terms such as cell division and chemical synaptic transmission (**Supplementary Figure 6B)**. The CRISPR deletion and GL_007 deletion lines exhibited significantly lower eigengene scores for the NPC-associated module (*blue*) at the NPC stage and significantly higher eigengene scores for the mature neuron-associated module (*turquoise*) at both the NPC and iMN stages **(Supplementary Figure 6C)**. These results suggest premature differentiation of NPCs. Furthermore, gene set enrichment analysis (GSEA) comparing deletion lines to nondeletion lines from the isogenic setting and each family across four differentiation stages revealed significant enrichment for GABAergic neuron and DNA replication-related terms, as well as for signaling pathways associated with PI3K-AKT, TGF-β, and Wnt **(Figure 3G, Supplementary Figure 6D, Supplementary Table 8).** Together, these findings suggest that the combined effect of the deletion and secondary variants lead to family-specific alterations in neurogenesis and associated signaling pathways.

### CRISPR activation of 16p12.1 genes reverses transcriptomic and neuronal defects

We previously found that the 16p12.1 genes exhibit diverse molecular functions and show limited connectivity within a brain-specific co-expression network, especially when compared to genes within the more penetrant 16p11.2 deletion^13, 45, 46^. Therefore, we sought to investigate the role of individual 16p12.1 genes in driving distinct functional changes across genetic backgrounds. We designed single-guide RNAs (sgRNAs) targeted to the promoters of *MOSMO*, *POLR3E*, and *UQCRC2* and used CRISPR activation (dCas9-VP64 and MS2-P65-HSF1-mediated CRISPRa^47^) to increase their expression in the deletion lines (see **Methods, Figure 4A, Supplementary Figure 7A)**. We selected these genes as they showed significantly reduced expression in the deletion samples across neuronal differentiation cell states **(Supplementary Figure 2A)**. Previous studies have also implicated these genes in cellular functions such as Sonic hedgehog (Shh) signaling, immune response, and mitochondrial homeostasis^48–50^. We performed CRISPRa experiments at the iPSC stage in three proband lines to assess the contribution of individual 16p12.1 genes to their more severe and varied cellular phenotypes and confirmed sustained overexpression of the targeted genes in NPCs, the cell state in which most cellular effects were observed in this study **(Supplementary Figure 7A)**. However, neurosphere formation failed for CRISPRa of *MOSMO* in P2C_079. This could be potentially due to a dosage-sensitive role of *MOSMO* in the genetic background of P2C_079 in regulating Shh signaling^51, 52^, which is known to influence neurosphere formation^53^.

**Figure 4.**
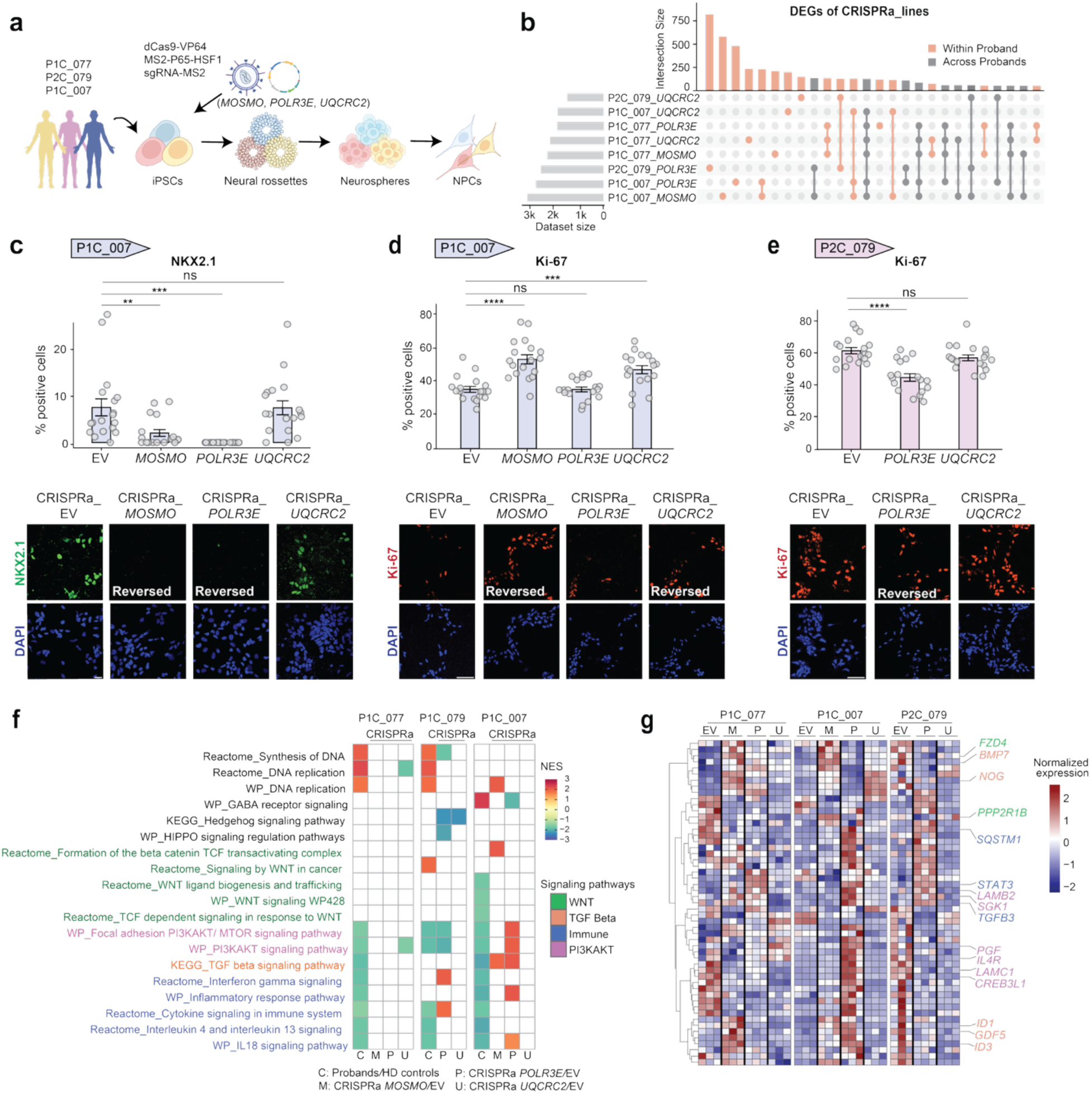
CRISPRa-mediated activation of 16p12.1 genes across probands. **(a)** Schematic of the CRISPRa approach (using dCas9-VP64 and MS2-P65-HSF1) in probands from the three families. **(b)** Upset plot shows overlaps among DEGs (log_2_|FC|≥0.5, padj<0.05) for each comparison (CRISPRa sgRNAs of deletion genes vs. empty sgRNA-MS2 vector) in the three proband lines. **(c)** Quantification and representative images of NKX2.1-positive cells with CRISPR activation of 16p12.1 genes in NPCs from P1C_007. Scale bars, 20 μm. **(d)** Quantification and representative images of Ki-67-positive cells with CRISPR activation of 16p12.1 genes in NPCs from P1C_007. Scale bars,70 μm. **(e)** Quantification and representative images of Ki-67-positive cells with CRISPR activation of 16p12.1 genes in NPCs from P2C_079. Scale bars, 70 μm. **(f)** Select pathways (q<0.05) enriched by GSEA across comparisons of different sets of DEGs (q<0.05) annotated in the Reactome, WP, and KEGG databases. Enrichment analysis with GSEA were made between DEGs obtained from deletion NPCs versus controls compared to CRISPRa versus EV controls for each proband. NES, normalized enrichment score; M, pathways derived from CRISPRa_*MOSMO* DEGs; P, pathways derived from CRISPRa_*POLR3E* DEGs; U, pathways derived from CRISPRa *UQCRC2* DEGs. White color fill indicates absence of terms in GSEA results. (**g)** Heatmap of normalized gene expression in selected pathways (colored in (**f**)) across CRISPRa lines, clustering using Ward’s D2. Color scale represents z-scores calculated from TPM within each group. EV, empty sgRNA-MS2 vector; M, CRISPRa of *MOSMO*; P, CRISPRa of *POLR3E*; U, CRISPRa of *UQCRC2*. Data are mean±s.e.m and one-way ANOVA followed by Dunnett’s post hoc test were used for **(c)**, **(d)**, and **(e)**. *p<0.05; **p<0.01; ***p<0.001; ****p<0.0001. n=3 independent experiments for each line, except for RNA-seq and downstream analyses where n=2 for iMNs and MNs of P1C_077, and MNs of HD_01. For **(c)**, **(d)**, and **(e),** n=6 images per experiment were quantified.

Using RNA-seq in NPCs, we identified DEGs comparing each CRISPRa line to its corresponding empty vector (EV) control. We found that most DEGs were unique to each proband, with overlaps primarily within the same proband, rather than across different probands, even when the same 16p12.1 deletion gene was activated **(Figure 4B, Supplementary Table 4)**. Furthermore, CRISPRa of each one of these genes did not change the expression of other 16p12.1 genes **(Supplementary Figure 7B, Supplementary Table 4)**. These findings suggest that CRISPRa-mediated restoration of 16p12.1 genes leads to outcomes modulated by the genomic background and that these deletion genes are not strongly interconnected, consistent with previous observations^46^.

We investigated whether CRISPR activation of individual 16p12.1 genes could reverse transcriptomic changes initially observed in probands with the deletion. By comparing DEGs obtained from CRISPRa experiments with those from probands, we identified 5,058 genes with opposing directions of gene expression change (i.e., reversed genes) **(Supplementary Figure 7C, Supplementary Table 4).** For example, restoring *POLR3E* reversed expression of 16 genes across all probands, including cytoskeleton-related (*PLS3* and *LIMA1*), cell adhesion (*PCDHA12*, *JAM2*, and *CD24*), and signaling transduction (*ZYX*, *S1PR1*, and *PDE1A*) genes **(Supplementary Figure 7D, Supplementary Table 4)**. Expression of specific phenotype-related genes were also reversed by activating distinct 16p12.1 genes. For example, the epilepsy risk gene *NNAT*^54^ was upregulated in P2C_079 and P1C_007, who both have epilepsy, and was reversed with CRISPRa of *POLR3E* in both probands. CRISPRa also reversed altered isoform usage; activating *MOSMO* restored isoforms of *MECP2* (ENST00000627864) and *EPS8L2* (ENST00000318562), genes that have been associated with autism and deafness, respectively, in P1C_077, who manifested these clinical features **(Supplementary Table 4)**^55, 56^.

Next, we investigated whether CRISPR activation of the 16p12.1 genes rescued cellular phenotypes. In P1C_007, overexpression of NKX2.1 was reversed with CRISPRa of *POLR3E* and *MOSMO*, while proliferation and apoptosis phenotypes were rescued by CRISPRa of *UQCRC2* and *MOSMO* **(Figure 4C-D, Supplementary Figure 8A**). Additionally, overexpression of *UQCRC2* lead to further enhanced premature differentiation of NPCs to neurons in this proband **(Supplementary Figure 8B)**. In P2C_079, CRISPRa of *POLR3E* rescued hyperproliferation phenotypes **(Figure 4E, Supplementary Figure 8C)**. These results underscore the direct role of 16p12.1 genes towards the neuronal defects observed in deletion carriers, with some functional effects observed among multiple genes and others being gene specific.

We further investigated the pathways altered by CRISPR activation of 16p12.1 genes and found enrichment for multiple signaling pathways such as TGF-β, PI3K-AKT, Wnt, and MAPK **(Supplementary Figure 8D, Supplementary Table 4).** Dysregulation of these pathways is known to disrupt cell-fate determination and cell-cycle progression^57–59^, as observed in the deletion lines. Therefore, we performed GSEA on the genes with altered expression from CRISPRa lines to assess the direction of change in associated signaling pathways compared to EV controls in proband lines **(Figure 4F, Supplementary Table 4).** We found that CRISPRa of *POLR3E* and *MOSMO* led to the restoration of TGF-β signaling, which corresponded to the rescue of the NKX2-1 expression level in P1C_007. Furthermore, activation of *POLR3E* also resulted in the upregulation of immune-related pathways, including interferon and cytokine signaling, which were downregulated in multiple deletion lines. Specific genes within these pathways, such as *STAT3* and *TGFB3* within immune pathways, and *FZD4*, *BMP7* and *NOG* within Wnt and TGF-β pathways, were also found to be dysregulated in these probands, potentially contributing to the reversal of proliferation and apoptosis phenotypes^60^ (**Figure 4G**). Together, these results suggest that each 16p12.1 gene modulates distinct signaling cascades in a genetic background-dependent manner, thereby contributing to variable functional trajectories across individuals.

### Connectivity of genes with secondary variants in PPI network

To further investigate the role of secondary variants and explain phenotypic differences across probands, we analyzed how genes with secondary variants whose expression was altered following CRISPRa of individual 16p12.1 genes in NPCs, were connected within a protein-protein interaction network^61^ **(Figure 5A, Supplementary Figure 9A, Supplementary Table 5)**. In the *POLR3E* and *MOSMO* CRISPRa lines, we observed a higher network connectivity for P1C_007 and P2C_079 compared to P1C_077, correlating with the presence of neuronal defects in NPCs of P1C_007 and P2C_079 but not P1C_077 **(Figure 5A).** These findings were further validated by analysis of PPI networks specifically within signaling pathways, which revealed higher network connectivity in P1C_007 and P2C_079 **(Figure 5B)**. Interestingly, in the *UQCRC2* CRISPRa lines, P1C_077 showed higher connectivity compared to P1C_007 and P2C_079 **(Supplementary Figure 9A-B).** The highly connected nodes within this network included proteins such as ADCY1, PDE4A, and GNG10, which are involved in cAMP signaling and G protein signaling pathways that modulate synaptic functions in mature neurons^62, 63^. This may explain the absence of cellular phenotypes in P1C_077 at the NPC stage. These findings indicate that the extent to which modifiers of individual 16p12.1 genes are connected within signaling pathways is a determinant of phenotypic variation across probands.

**Figure 5.**
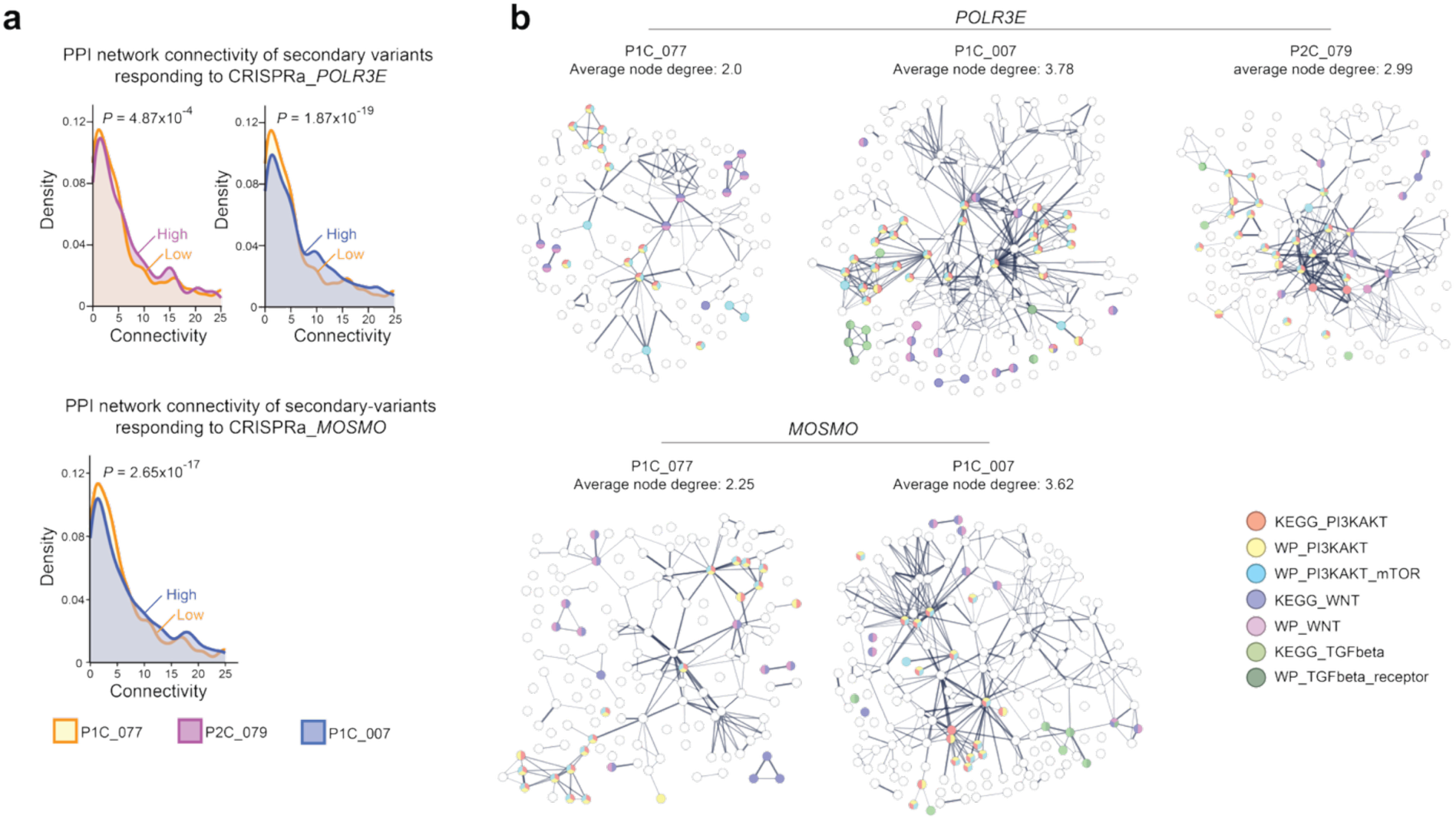
Connectivity of secondary variants across probands in response to CRISPR activation of 16p12.1 genes. **(a)** Density plots show the connectivity (node degree, i.e., the number of connections a node has to other nodes in the network) of *POLR3E* and *MOSMO* CRISPRa DEGs (log₂|FC|>0.5, padj< 0.05) that overlap with secondary variants, as measured using the STRING database. Anderson-Darling k-sample test was used to calculate p values. **(b)** Visualization of the PPI network of DEGs (log₂|FC|≥0.5, padj<0.05) that overlap with secondary variants and were restored by CRISPRa of *POLR3E* in P1C_077 (top left), P1C_007 (top middle), and P2C_079 (top right), as well as by CRISPRa of *MOSMO* in P1C_077 (bottom left) and P1C_007 (bottom right). DEGs shown were selected based on their involvement in signaling pathways annotated in the Reactome database. p-value <10^-16^ for all networks. Protein nodes involved in specific pathways are color-coded based on annotations in the STRING database. Average node degrees and PPI enrichment p values were also calculated using the STRING database. n=3 independent experiments for each line.

### Modulation of FOXG1 and JUN mediates phenotypic variability

We sought to identify functional convergence across samples through which the deletion confers disease susceptibility, modulation of which by secondary variants lead to diverse outcomes. As the network of genes within signaling pathways typically converge on TFs^64^, we sought to identify key TFs that regulate downstream effects in the deletion lines. We found that the sequences within Diffpeak regions across the deletion NPCs were enriched for binding motifs of TFs involved in developmental processes, such as FOS, HOXD13 and NR4A1^65–67^ **(Supplementary Table 6)**. Furthermore, co-regulatory networks constructed from these TFs whose binding motifs showed the most significant enrichment in Diffpeak regions across the deletion lines identified FOXG1 and JUN as two of the top-ranked TFs using ChEA3 **(Figure 6A, Supplementary Table 6**, see **Methods).** Both of these TFs have been previously implicated in multiple signaling pathways, including Wnt, BMP, and JNK^68–70^. Furthermore, we observed altered expression of FOXG1 and JUN across the deletion lines compared to controls, along with changes in both expression and chromatin accessibility of genes within their regulatory networks. These included genes such as *HES1*, *KLF6*, *DLX2*, and *FZD8,* which are known to play roles in neural processes such as inhibitory neuron production and NPC proliferation^71–74^ (**Figure 6A, Supplementary Fig. 10A-B, Supplementary Table 6).** Therefore, we hypothesized that the 16p12.1 deletion affects regulatory hubs within signaling pathway interaction networks, and that modulating the expression of individual 16p12.1 genes would, in turn, alter the expression of these hub genes, such as FOXG1 and JUN, as well as their connected genes, leading to changes in cellular phenotypes (**Figure 7**). To test this, we further investigated the roles of FOXG1 and JUN towards the observed cellular dysregulations in the deletion lines. Since altered activity of the TFs reflects perturbations across multiple associated signaling pathways, we did not expect a direct one-to-one correspondence between specific TF activity and cellular phenotypes. We found that the expression levels of FOXG1 and JUN varied across probands, and these levels were differentially modulated with CRISPR activation of different 16p12.1 genes **(Figure 6B-C)**. Changes in FOXG1 and JUN expression in the P1C_007 CRISPRa lines, and changes in JUN expression in the P2C_079 lines, were observed alongside the reversal of cellular phenotypes upon CRISPR activation **(Figure 4**, **Figure 6D)**. Furthermore, we found that the expression of genes within their regulatory networks, including *DLX2* and *KLF6*, mirrored changes in *FOXG1* and *JUN* expression across CRISPRa lines **(Figure 6E)**. These findings suggest that key TFs and the regulatory network within individual 16p12.1 gene-associated signaling pathways mediate the phenotypic trajectories of the deletion (**Figure 7**).

**Figure 6.**
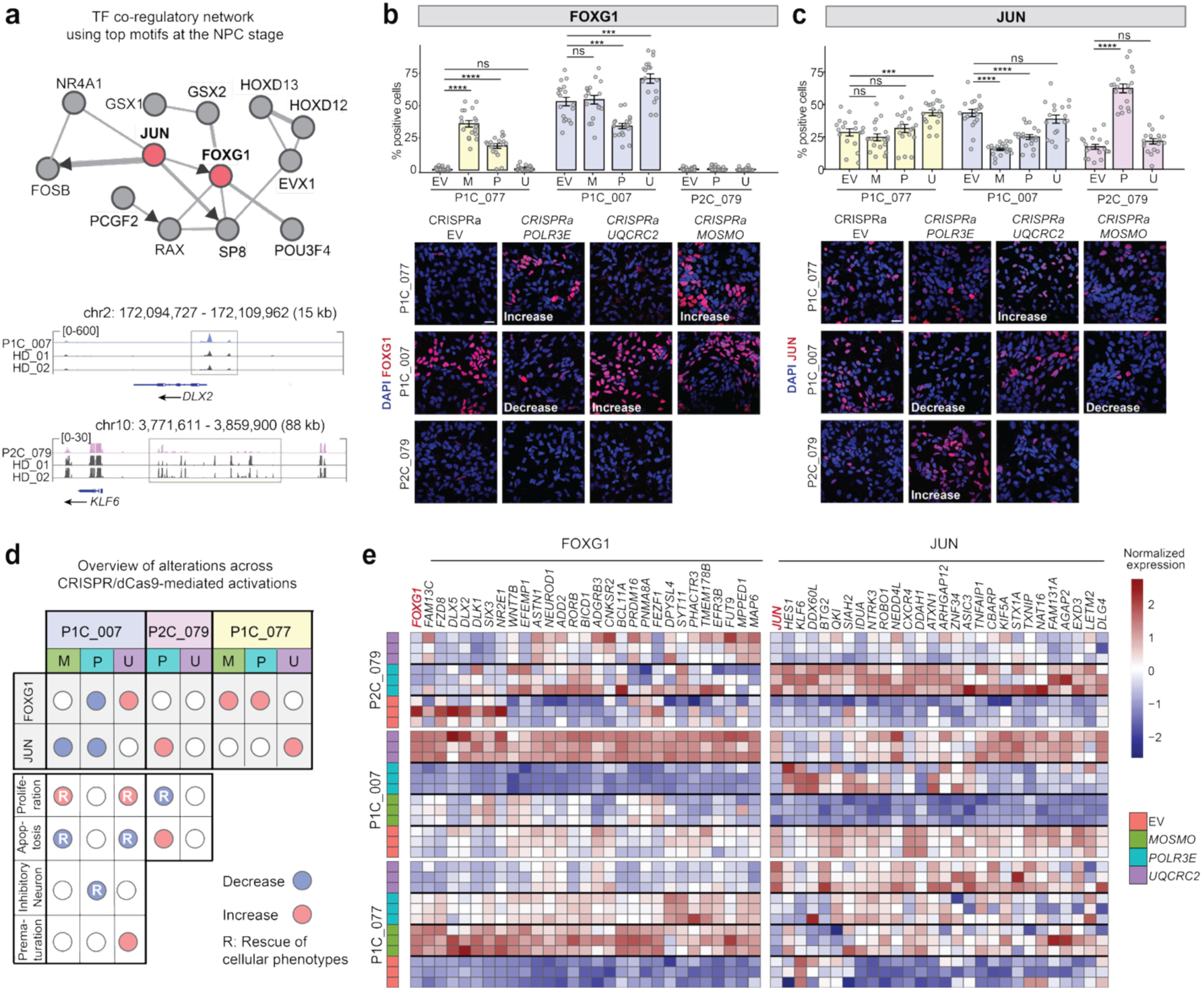
Modulation of gene regulatory hubs corresponding with variable phenotypes. **(a)** TF co-regulatory network constructed by ChEA3 using TFs whose binding motifs were most significantly enriched in ATAC-seq Diffpeak regions from comparison of deletion lines vs. healthy donor lines at the NPC stage. Motif lists are provided in **Supplementary Table 6** (see **Methods**). The panel below depicts IGV snapshots of ATAC-seq peaks showing increased (*DLX2*) or decreased (*KLF6*) chromatin accessibility for select genes in the regulatory network of FOXG1 and JUN, respectively. **(b)** Quantification and representative images of FOXG1-positive NPCs in CRISPR activated lines from P1C_077, P1C_007, and P2C_079. Scale bars, 20 μm. EV, empty sgRNA-MS2 vector; M, CRISPRa of *MOSMO*; P, CRISPRa of *POLR3E*; U, CRISPRa of *UQCRC2*. **(c)** Quantification and representative images of JUN-positive NPCs in CRISPR activated lines from P1C_077, P1C_007, and P2C_079. Scale bars, 20 μm. **(d)** Overview of CRISPRa results at the NPC stage. Columns represent CRISPRa lines from the three probands; rows indicate the direction of change based on phenotype quantification. Empty bubbles denote no detectable change. Rescue of cellular phenotype refers to the reversal of a cellular phenotype observed in the proband lines (compared to healthy donor controls). The phenotypes of proliferation, apoptosis, inhibitory neuron, and premature maturation were, respectively, measured with EdU, TUNEL assay, VGAT, NKX2.1 and TUBB3 positive cells. **(e)** Representative heatmap of normalized expression of genes within the regulatory networks (identified using ChEA3) of FOXG1 (left) and JUN (right) across the CRISPRa lines. Color scale represents the z-scores calculated from TPM within each group. Data are mean±s.e.m and one-way ANOVA followed by Dunnett’s post hoc test for **(b)** and **(c)**. *p<0.05; **p<0.01; ***p<0.001; ****p<0.0001. n=3 independent experiments for each line. 6 images per experiment were quantified for **(b)** and **(c)**.

**Figure 7.**
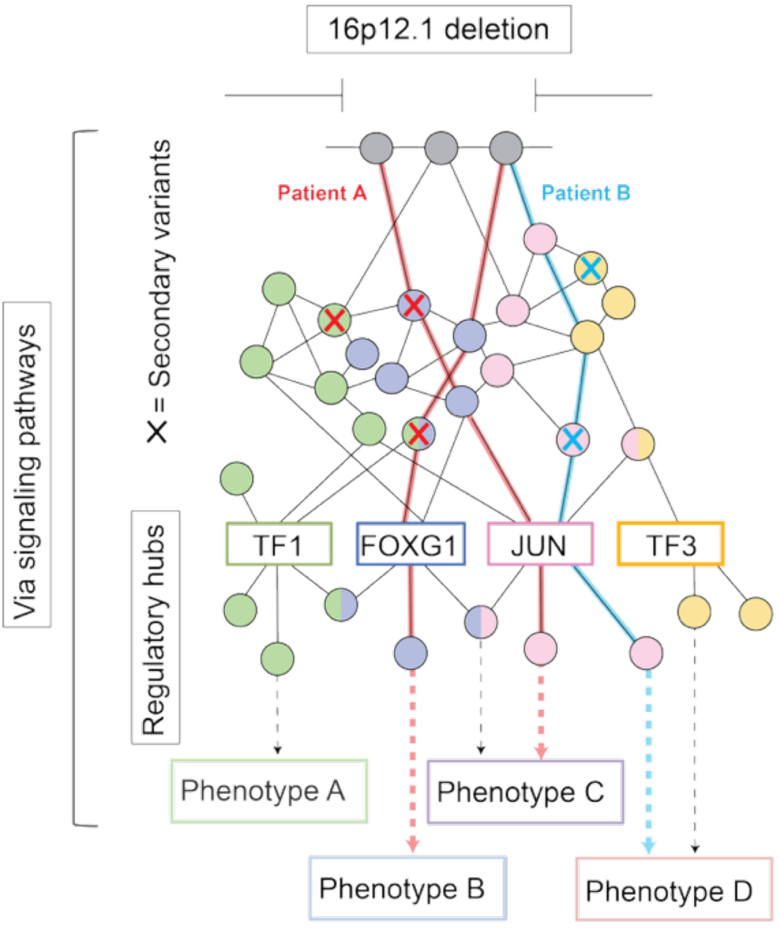
Conceptual model illustrating how the interplay between the 16p12.1 deletion and secondary variants contributes to variable expressivity. The interactive effects are mediated through regulatory hubs, such as TFs (e.g., FOXG1, JUN), within signaling pathways. The network is colored for the TF regulatory hub, and two-toned colors indicate genes shared among multiple pathways. Patient-specific secondary variants or combinations of variants lead to distinct phenotypic trajectories, shown by the red or blue connecting lines traversing distinct genetic pathways.

## DISCUSSION

Our study provides a framework to understand how variable expressivity in neurodevelopmental disorders arises from the complex interplay between disease-associated variants and secondary variants in the genetic background. Using the 16p12.1 deletion as a model, we found that primary and secondary variants jointly modulate key signaling pathways and shape distinct phenotypic trajectories through transcription factors within gene regulatory networks (**Figure 7**). Several themes have emerged from our work, contributing to the emerging picture that network-level genetic interactions underlie variable expressivity in complex genetic disorders.

Previous studies have linked CNVs to changes in chromatin accessibility and gene expression in neurodevelopmental disorders^75, 76^. Here, we observed potential nonadditive effects of primary and secondary variants on both chromatin accessibility and gene expression. Notably, even the ostensibly healthy control used to generate the CRISPR deletion line did not fully control the genetic background effects and therefore cannot isolate the effect of the deletion alone. The presence of secondary variants interacting with the deletion highlights the need to carefully account for genetic background effects in future studies, even when using isogenic models, as introducing the same variant into distinct lines may yield different results when characterizing variant function.

Although observed within specific genetic backgrounds, the 16p12.1 deletion led to defects in inhibitory neuron production, abnormal cell proliferation and apoptosis, and premature neuronal differentiation. These findings reflect disruptions in excitatory/inhibitory balance, neurogenesis dynamics, resulting in altered cell type proportions during brain development, a hallmark of multiple neurodevelopmental disorders including ASD, schizophrenia and intellectual disability^19, 41, 77^. Furthermore, CRISPRa-based rescue of 16p12.1 genes supports our previous findings^46^ that each gene within this region functions independently, as rescue of different genes reversed distinct sets of dysregulated genes and cellular phenotypes without affecting the expression of other deleted genes. Future work using this framework could incorporate CRISPR interference-based gene knockdown and be extended to other variably expressive CNVs, such as those at 1q21.1, 15q13.3, 16p11.2, and 22q11.2, to link genes within these regions to cellular phenotypes across the diverse genetic backgrounds of individual patients.

We observed alterations across multiple signaling pathways, including Wnt, TGF-β, PI3K-AKT, and immune-related pathways, several of which have been implicated in neurodevelopmental disorders^58, 78–81^. These findings reflect the extensive crosstalk and network-level interconnectivity among these pathways^78^. Furthermore, we found that the severity of phenotypic outcomes correlated with the PPI connectivity of secondary variants in specific signaling pathways. In addition, consistent with prior studies identifying convergent mechanisms underlying heterogeneity in autism^82^, we found differential modulation of transcription factors such as FOXG1 and JUN, which serve as regulatory hubs across deletion lines with distinct genetic backgrounds. TFs have been shown to be key regulators of cellular differentiation, and their dysregulation has been linked to developmental disorders, including craniofacial anomalies^83^. Together, these results underscore the importance of examining both inter-individual variability and the convergent mechanisms that emerge across diverse genetic contexts.

While our study provides key insights into the functional impact of secondary variants, several aspects could be further refined or expanded in future work. *First*, while we observed strong associations between secondary variants and changes in gene expression and chromatin accessibility, the small sample size limits our ability to infer causality without further experimental validation. High-throughput approaches such as Perturb-seq and CROP-seq could help confirm these interactions and elucidate how they modulate signaling pathways^84, 85^. *Second*, we used 2D iPSC differentiation combined with bulk RNA-seq, which may overlook cell type– specific effects, particularly in neuronal subtypes and non-neuronal brain cells relevant to neurodevelopmental disorders^38^. Future work could leverage advanced models such as neural organoids, assembloids, chimeroids, or other emerging platforms, which provide greater cellular diversity and spatial resolution^86^. Overall, our findings lay the groundwork for future functional studies to explore genotype–phenotype relationships, inter-individual variability, and potential personalized therapeutic strategies.

In summary, our study provides a framework for understanding how the combined effects of disease-associated variants and genetic background drive variable expressivity in neurodevelopmental disorders. By uncovering both shared and background-specific impacts on gene regulation and signaling pathways, we underscore the importance of accounting for genetic context in functional studies and in developing personalized therapeutic strategies.

## MATERIALS AND METHODS

### Patient recruitment and clinical phenotype analysis

Patient recruitment and clinical phenotyping were performed as previously described^14^. Families were recruited through clinics by obtaining de-identified samples and clinical data based on an approved protocol (IRB #STUDY00017269). Detailed medical histories, including clinician-reported, guardian-reported (for children), or self-reported (for adults), were collected, and standardized questionnaires were administered to assess developmental phenotypes in children and psychiatric features in adults. Questionnaires for children assessed neuropsychiatric and developmental features, anthropometric measurements, congenital anomalies in multiple organ systems, and family history of medical or psychiatric conditions. For assessing quantitative phenotypes, we conducted quantitative assessment using the Hansen Research Services Matrix Adaptive Test (HRS-MAT)^87^ to evaluate nonverbal IQ and the Social Responsiveness Scale (SRS)^88^ to assess autism-related social behavior. HRS-MAT was self-administered by participants via an online platform. SRS was delivered through a RedCap-based survey platform maintained by the Geisinger Autism and Developmental Medicine Institute. For participants aged 18 or older, the SRS was self-reported; for those under 18, responses were provided by parents or guardians. Body Mass Index (BMI) and head circumference were obtained either from medical records or self/guardian-reports. BMI was calculated from height and weight data obtained from medical records or self/guardian-reports where necessary. Both BMI and head circumference were converted to age- and sex-adjusted z-scores. For the assessment of developmental milestones, we followed the CDC guidelines^89^. For detailed phenotypic data and related information, please contact the corresponding author.

### Induced pluripotent stem cell reprogramming and maintenance

Induced pluripotent stem cells (iPSCs) were reprogrammed from peripheral blood mononuclear cells using the Cytotune-iPS 2.0 Sendai Programing kit, including the four Yamanaka reprogramming factors POU5F1 (OCT4), SOX2, KLF4, and MYC (ThermoFisher Scientific) as described previously^90^, and validated by flow cytometry (EPCAM, TRA-1-81, SSEA4 and CD9) and immunofluorescence (NANOG, OCT4, SOX2 and SSEA4). HD_01 and HD_02 were derived from CD34+ cord blood from healthy male and female donors, respectively (BID00274 and BID00271, NINDS). These lines were grown in mTESR1 (#85850 StemCell Technologies) or mTESR plus medium (#100-0276 StemCell Technologies), supplemented with 1% Penicillin/Streptomycin (# P4333 Sigma-Aldrich) on dishes coated with Geltrex (#A1413302 Gibco), and were passaged using 0.5mM EDTA (#AM9260G Invitrogen) or ReLeSR (#100-0483, StemCell Technologies), with Rock inhibitor (Y-27632) to improve cell survival (# 72304 StemCell Technologies). Cells were incubated at 37⁰C and 5% CO_2_. Mycoplasma tests were performed on iPSCs and NPCs using mycoplasma PCR detection kit (#MP0035, Sigma-Aldrich).

### Generation of CRISPR/Cas9-mediated 16p12.1 deletion iPSC line

The sequences of two sgRNAs are: (1) TCGGTGCTTAGGATCAGCCT, (2) GCCACTAGCTGACATGGTTG. Two separate vectors were used for delivering the sgRNAs: pSpCas9(BB)-2A-Puro (PX459) V2.0 was a gift from Feng Zhang (Addgene plasmid # 62988) and pGH020_sgRNA_G418-GFP was a gift from Michael Bassik (Addgene plasmid # 85405). Two vectors with sgRNAs were transfected into the HD_01 line using Lipofectamine stem transfection reagent (# STEM00001, Invitrogen), and the CRISPR control line was generated via transfection of empty vectors. Puromycin selection (0.5ng/ml) was performed 24 hours after transfection and lasted an additional 24 hours before the selection medium was replaced with mTESR1 medium. The cells recovered for several days before being diluted and split into 96-well plates manually to isolate single-cell clones. Each clone was cultured for two weeks and genotyped by the following two set of primers: WT-F: GACTTCCTCCACATCTTCCTCTA, WT-R: TCAAATAGAGGGGCAGGAGC (546bp); Deletion-F: TCCTCAGACTCAATAATTGCCA, Deletion-R: TGACCTTTACTCTGTGACATTGC (942bp). The desired PCR products were purified, and sanger sequencing was used to confirm that the correct sequence was present. For further conformation, genomic DNA of the selected clones was extracted by GenElute Mammalian Genomic DNA Miniprep kits (#G1N350, Sigma-Aldrich) and PureLink Genomic DNA Mini kit (#K182001, Invitrogen) for SNP-array.

### RNA isolation and RT-qPCR

Total RNA was isolated using the TRIzol reagent (#15596026, Invitrogen) and PureLink RNA mini kit (#12183018A, invitrogen), following a modified version of this kit’s protocol. Cells were collected directly in TRIzol and frozen until RNA extraction. Chloroform was added to the TRIzol in a 1:5 ratio, shaken vigorously for 30 seconds, then incubated for 3 minutes before being centrifuged at 12,500 g for 15 minutes at 4°C. The upper phase was collected in a fresh tube and an equal amount of ethanol was added before vortexing. The mixed reagent was transferred to a spin column, and washing and recovery steps proceeded as described in the kit protocol. Isolated RNA was treated with TURBO DNase (catalog #AM1907, Thermo Fisher scientific, MA, USA). RT-qPCR was performed in three independent experiments, each with three replicates, for each sample. 1000ng RNA was used in reverse transcription reaction by qScript cDNA Synthesis Kit (Quantabio, catalog#95047-100). The cDNA product was diluted 100 times and 2ul cDNA was used in 10ul RT-qPCR reaction, using PowerTrack SYBR Green Master Mix (catalog#A46109, Applied Biosystems). RT-qPCR was performed using the QuantStudio 3 (Applied Biosystems) through the following steps: initial denaturation step at 95 °C for 2 min, followed by 40 cycles of denaturation at 95 °C for 15 s and annealing/elongation at 60°C for 1 min. A melt curve analysis was conducted at the end of the amplification. The results were analyzed using QuantStudio Design & Analysis software. The following primers were used: GAPDH-F: ATGGGGAAGGTGAAGGTCGG, GAPDH-R: TGACGGTGCCATGGAATTTG; POLR3E-F: GGAGCAGATTGCGCTGAA, POLR3E-R: TTACTGGTGGTCTGGGAAGA; UQCRC2-F: TTCAGCAATTTAGGAACCACCC; UQCRC2-R: GGTCACACTTAATTTGCCACCAA; MOSMO-F: CTGTCACATGTGGTTTGCTGG; MOSMO-R: GGGCAGCCATACAGAAAAGGA.

### Neural conversion of iPSCs to NPCs

The neural conversion of iPSCs was performed as previously described with modifications^18^. On day 0, iPSCs were treated with Collagenase Type IV (StemCell Technologies #07909). Then, cells were scraped and transferred to an uncoated 60mm dish with mTESR1 media supplied with 10uM Y-27632 for embryoid body (EB) formation. After 48 hours in suspension, mTESR1 media was switched to neural differentiation media (N2 media) containing DMEM/F-12 (catalog# 10565018, Gibco) with 1% N-2 Supplement (catalog#17502001, Gibco) and 1% MEM Nonessential amino acids (catalog#11140050, Gibco), 2ug/ml Heparin (catalog#07980, StemCell Technologies), 1% Penicillin/Streptomycin (catalog#P4333, Sigma), 5uM SB431542 (catalog#72232, StemCell Technologies) and 0.25uM LDN193189 (catalog#72147, stem cell technologies). The EBs were cultured in neural differentiation media for 4 days and the media was changed every two days. On day 6, EBs were seeded on Geltrex-coated plates for 8 days to form rosettes. From days 6-8, rosettes were cultured in N2 media without Dual-SMAD inhibitors, and from days 8-14, they were cultured in N2B27 media containing 1:1 DMEM/F-12 and Neurobasal Medium (catalog#21103049, Gibco) with GlutaMax supplement (catalog#35050061, Gibco), 1% N-2 Supplement, 2% B-27 supplement minus vitamin A (catalog#12587010, Gibco), 1% MEM Nonessential amino acids, 2ug/ml Heparin, 1% Penicillin/Streptomycin. On day 14, rosettes were treated with Collagenase Type IV and then transferred to uncoated 60mm dish with N2B27 media for neurosphere formation in suspension culture for 12 days. On day 26, neurospheres were dissociated by Accutase (catalog#A1110501, Gibco) then plated on Geltrex-coated plates. Neural progenitor cells were maintained in Stemdiff neural progenitor medium (catalog#05833, StemCell Technologies) and passaged 1:2-1:3 using Accutase. This protocol is expected to create a population of NPCs which express SOX2 and NESTIN, with the majority of cells expressing Pax6.

### Neuronal differentiation of NPCs to neurons

Plates were prepared for neuronal differentiation with Poly-D-Lysine (PDL, catalog#A3890401, Gibco) and Laminin (catalog#L2020, Sigma-Aldrich). PDL was diluted 1:1 with PBS to a concentration of 50ug/ml and left in the wells for 1 hour at room temperature. PDL was aspirated and the well was washed 4-5 times with sterile DI water and then allowed to dry for 15 minutes. Laminin was diluted to 20ug/ml in chilled dPBS and added to the wells. After characterization, NPCs were plated 800,000 cells/per well on the 6-well plates. The next day, 0.2uM Compound E (catalog#73952, StemCell Technologies) was added to the media. In the first two weeks, neuronal differentiation media was used, containing Neurobasal medium, 1% GlutaMAX, 1% Penicillin/Streptomycin, 1% N-2 Supplement, 2% B-27 supplement (#17504044, Gibco), 0.2% MycoZap Plus-PR (catalog#195263, LONZA) supplied with 20ng/ml BDNF (#450-02-1mg, Pepro Tech), 20ng/ml GDNF (450-10-1mg, Pepro Tech), 200uM Ascorbic acid(#72132, StemCell Technologies), 1uM cAMP (#A9501) and 1ug/ml Laminin. In the last two weeks, the neuronal differentiation media was switched to BrainPhys Neuronal medium (catalog#05790, StemCell Techonologies), supplemented with 40uM 5-Fluoro-2′-deoxyuridine (FUDR) (catalog#F0503, Sigma-Aldrich) to kill mitotic cells. Half media changes were performed every 3 days.

### Cell proliferation and apoptosis

Proliferation and apoptosis assays were adapted from a previous study^18^. On day 0, NPCs at passage 6 were seeded at 1x10^5^ on 18mm diameter coverslips coated in PDL (1:1 in PBS) and Geltrex (1:100 in DMEM/F12) on 12-well cell culture plates in neuronal differentiation medium containing Neurobasal media, 1% N-2 supplement, 2% B-27 supplement minus vitamin A, 1% GlutaMax, 1% Penicillin/Streptomycin, and were incubated overnight. On the morning of day 1, neural progenitor cells were treated with 10uM 5-ethynyl-2’-deoxyuridine (EdU) (catalogC10337, Invitrogen) for four hours and before the media was replaced with fresh medium without Edu. Coverslips were collected and fixed at three timepoints, of no chase (0 hours), 24-hour chase, and 48-hour chase. The Edu label was detected via Click-iT chemistry following manufacturers’ instructions. At the 24-hour chase timepoint, coverslips were also stained for Ki-67 markers (1:200 catalog#12202, Cell Signaling TECHNOLOGY). On day 5, TUNEL assay for apoptosis detection was performed using Click-iT Plus TUNEL Assay kit (catalog#C10617, Invitrogen).

### iPSC and NPC immunostaining

NPCs at passage 6 or iPSC clones were seeded on coverslips coated in PDL (1:1 in PBS) and Geltrex (1:100 in DMEM/F12) in 6-well plates. Media was aspirated and cells were washed using PBS three times. Cells were fixed in 4% paraformaldehyde (PFA) (catalog#158127, Sigma-Aldrich) for 15 minutes, washed three times with PBS, then permeabilized in 0.2% Triton X-100 (catalog#X100, Sigma-Aldrich) in PBS for 15 minutes. Blocking was done with a solution of 2% BSA (catalog#A7906, Sigma-Aldrich), 0.2% Triton X-100, and 5% goat serum (catalog# G9023, Sigma-Aldrich) in PBS for 1 hour at room temperature or overnight at 4 °C. The coverslips were incubated with primary antibodies at room temperature for 2 hours or overnight in the 4°C. Coverslips were washed with PBS three times for 5 minutes each, then incubated with the secondary antibody for 2 hours at room temperature. Coverslips were again washed in PBS three times and mounted using DAPI antifade mounting reagent. All antibodies were diluted in blocking buffer. Antibodies: SOX2 (1:200, catalog#3579S Cell Signaling Technology); NESTIN (1:800, catalog#33475S Cell Signaling Technology); NKX2.1 (1:200, catalog#MAB5460, Millipore Sigma); PAX6 (1:800, catalog#60433S, Cell Signaling Technology); TUBB3 (1:400, catalog#5568T, Cell Signaling Technology); FOXG1 (1:400, calalog#29642S, Cell Signaling Technology); c-JUN (1:200, catalog#9165T, Cell Signaling Technology); Ki-67 (1:200, catalog#9129T, Cell signaling Technology); SSEA4 (1:200, catalog#4755T, Cell Signaling Technology); NANOG (1:500, catalog#4893T, Cell signaling Technology); OCT4 (1:200, catalog#2750S, Cell Signaling Technology); Anti-mouse IgG (H+L), F(ab’)2 Fragment (1:1000,catalog #4408S, Cell Signaling Technology); Anti-rabbit IgG (H+L), F(ab’)2 Fragment (1:1000, catalog#8889S, Cell Signaling Technology).

### Neuron immunostaining

Neurons were differentiated for 28 days then fixed with 20% PFA added directly to media to reach a final concentration of 4% and incubated at room temperature for 15 minutes. PFA was removed and cells were washed in PBS once for 5 minutes. 0.2% Triton X-100 in PBS was added and incubated room temperature for 15 minutes to permeabilize. Triton X-100 was removed, and cells were blocked at room temperature for 1 hour in 2% BSA, 0.2% Triton X-100, and 5% goat serum in PBS. The coverslips were incubated with primary antibodies at room temperature for 2 hours or overnight in the 4°C refrigerator. Coverslips were washed with PBS two times for 5 minutes each, then incubated with the secondary antibody for 2 hours at room temperature. Coverslips were again washed in PBS 2 times for 5 minutes each, then mounted using DAPI antifade mounting reagent. All antibodies were diluted in blocking buffer. Antibodies: MAP2 (1:500, catalog#4542S, Cell Signaling Technology); VGAT (1:250, Synaptic Systems); VGLUT1 (1:500, Synaptic Systems). NeuN (1:400, catalog#94403S, Cell Signaling Technology); Anti-mouse IgG (H+L), F(ab’)2 Fragment (1:1000, catalog #4408S, Cell signaling Technology); Anti-rabbit IgG (H+L), F(ab’)2 Fragment (1:1000, catalog#8889S, Cell signaling Technology).

### Lentivirus packaging

HEK293T cells were grown in media containing DMEM (catalog#D6429, Sigma-Aldrich) and 10% FBS (catalog#F2442, Sigma-Aldrich), 1% Penicillin/Streptomycin. On day 0, cells were seeded 18-24 hours prior to transfection to reach 80%-95% confluency on the day of transfection. On day 1, fresh media was added 30 minutes prior to transfection. Transfection was performed using TransIT-Lenti transfection Reagent (catalog#MIR6604, Mirus Bio), Opti-MEM (catalog#31985070, Gibo), and LV-MAX Lentiviral Packaging Mix (catalog#A43237, Thermo Fisher). On day 2, fresh media was added with ViralBoost Reagent (catalog#VB100, ALSTEM CELL ADVANCEMENTS). On day 3, the supernatant was collected and centrifuged at room temperature for 10 min at 300g to pellet debris and then the supernatant was passed through a 0.45um filter. Lentivirus Precipitation Solution (catalog#VC100, ALSTEM CELL ADVANCEMENTS) was added to supernatant and a ratio of 1:4, mixed thoroughly, and incubated overnight at 4°C, and then centrifuged for 30 min at 1500g at 4°C. The virus pellet was diluted using mTESR1 medium and stored at -80°C.

### RNA-seq sample processing and analysis

Each iPSC line was grown in three replicates, which were differentiated separately. RNA was extracted from iPSCs, neural progenitor cells (NPCs; passage 5), and neurons differentiated from NPCs at day 10 (iMNs) and day 28 (MNs). Only samples with RNA integrity number (RIN) ≥ 6.0, as measured by the Agilent TapeStation 4200 (Agilent Technologies), and purity (A260/280 ratio >1.8) were subjected to RNA sequencing. RNA sequencing libraries were prepared using the NEBNext Ultra II RNA library Prep kit (#E7770S, NEB) for Illumina according to the manufacturer’s instructions. 150 base pair (bp) paired-end sequencing were performed using the Illumina NovaSeq 6000 platform by Genewiz from Azenta Life Sciences (South Plainfield, NJ) for 30 million reads. Trimmomatic v0.39^91^ were used to trim adapters and to remove low-quality reads (leading:3, trailing:3, slidingwindow:4:15, and minlen:36 parameters). Kallisto v.050.0^92^ with n=100 bootstrap samples was used to quantify the abundances of transcripts and hg38 cdna was used to build the Kallisto index. Differential gene expression was analyzed using the DEseq2 v1.44 package in R v.4.3.1^93^. Batch effects were corrected by Combat-seq using sva v.3.35.2 package in R v.4.3.1^94^.

### Detection of alternative isoform usage

Alternative isoform usage was analyzed using the IsoformSwitchAnalyzeR v.2.4.0.^95^ package in R. Abundance files from Kallisto were input using the importIsoformExpression function. Genes or isoforms under the cutoff were removed by the prefilter function at default settings. Isoform switches were identified by isoformSwitchTestDEXseq function. To identify alternative isoform usage events independent of overall gene expression changes, isoforms belonging to genes that were also identified as DEGs were excluded.

### Weighed gene co-expression network analysis (WGCNA)

WGCNA was performed using the WGCNA 1.72 package in R^96^. For WGCNA analysis using the transcriptome data of NPC, iMN, and MN in Supplementary Figure 6A-B, first, raw counts were normalized by variance stabilizing transformation (VST) from DEseq2 1.44 package in R. The top 10,000 variable genes were extracted by median absolute deviation (MAD). The power (β) was chosen by pickSoftThreshold function with an R.sq cutoff of 0.8. Signed hybrid co-expression network (TOMType = “unsigned”, minModuleSize = 100, mergeCutHeight =0.25) was built by a_ij_ =[cor(x_i_, x_j_)]^β^, for cor(x_i_, x_j_) >0; a_ij_ = 0 for cor(x_i_, x_j_)≤0. Module eigengene was calculated as the first principal component to represent the expression profiles in each model. Pearson’s correlation coefficient was used to assess correlations between modules and traits.

### Whole genome sequencing (WGS) sample processing and analysis

DNA was isolated from blood samples from 22 participants and Illumina TruSeq DNA PCR-free libraries (San Diego, CA, USA) were constructed for 150bp paired-end whole-genome sequencing using Illumina HiSeq X by Macrogen Labs (Rockville, MD, USA). DNA from five samples (HD_01, HD_02, CRISPR deletion, P2C_079, and FNC_079) were isolated from iPSC cell lines using GenElute Mammalian Genomic DNA miniprep kits (#G1N79-1KT SIGMA). The genomic DNA sample was broken into short fragments. These DNA fragments were then end-polished, A-tailed, and ligated with full-length adapters for Illumina sequencing before further size selection. PCR amplification was then conducted unless specified as PCR-free. Purification was performed through the AMPure XP system. The resulting library was assessed on the Agilent Fragment Analyzer System and quantified to 1.5nM through Qubit and qPCR. Whole genome sequencing was conducted using Illumina Novaseq X by Novogene (Durham, NC, USA). Samples were sequenced at an average 37.9X coverage, with an average of 810M reads per sample and 98.0% of reads mapping to the human genome. We followed the GATK Best Practices pipeline^97^ to identify single nucleotide variants (SNVs) and small indels using GATK v.4.5.0. GATK’s MarkIlluminaAdapters was used to first mark adapter sequences and then align the reads to the GRCh38 reference genome using BWA v.0.7.173^98^. Duplicate reads were removed using the Picard implementation of MarkDuplicates and base quality score recalibration was performed using the BaseRecalibrator and ApplyBQSR. HaplotypeCaller was used to call variants for each sample, merge all variants into a single GVCF, and perform joint genotyping using GenotypeGVCFs. We then used the Python API for Hail and Ensembl Variant Effect Predictor (VEP) v.109^99^ to annotate variants. After splitting multi-allelic sites, variants were filtered for those with (i) ≥90% call rate, (ii) Hardy-Weinberg equilibrium p value ≥10-15, (iii) read depth ≥8, and (iv) allele balance ≥0.2. Variants were further filtered for those with a gnomAD v.2.17^100^ frequency <0.1%. We annotated variant consequences based on transcript consequences annotated by VEP and filtered for variants that were LOF (“transcript_ablation”, “stop_gained”, “frameshift_variant”, “stop_lost”, “start_lost”), missense (“missense_variant”), splice LOF (“splice_acceptor_variant”, “splice_donor_variant”), splice (“splice_donor_5th_base_variant”, “splice_region_variant”, “splice_donor_region_variant”, “splice_polypyrimidine_tract_variant”), upstream (“upstream_gene_variant”), downstream (“downstream_gene_variant”), 5’ UTR (“5_prime_UTR_variant”), 3’ UTR (“3_prime_UTR_variant”), or intronic (“intron_variant”). We used dbNSFP8–10 v.4 annotations to filter missense variants for those predicted to be deleterious by at least five of nine selected tools (SIFT, LRT, FATHMM, PROVEAN, MetaSVM, MetaLR, PrimateAI, DEOGEN2, and MutationAssessor). Variants were finally filtered for those private to related individuals to account for any technical differences between our data and gnomAD. STRs were initially called using GangSTR v2.5.0^101^, followed by quality control and filtering with DumpSTR v6.0.1. STRs with low-quality calls and those with read depths outside the range of 20 to 1000 were excluded using the --gangstr-min-call-DP and --gangstr-max-call-DP parameters. Additional quality control measures included filtering out all reads except spanning and bounding (--gangstr-filter-spanbound-only) and filtering regions with poorly estimated confidence intervals (--gangstr-filter-badCI). Individual VCF files were then merged into a multi-sample VCF using MergeSTR v.6.0.1. To identify rare STRs, loci with an allele frequency of <0.1% in EnsembleTR (https://github.com/gymrek-lab/EnsembleTR) were initially retained. Summary statistics were calculated using StatSTR v.6.0.1^102^. For frequency >0.1%, only loci with repeat lengths exceeding two standard deviations above the population mean were kept. Additionally, rare STRs defined as those present in fewer than eight individuals in the iPSC cohort were selected for downstream analysis. Final STR annotations were performed using ANNOVAR^103^. CNVs were called using CNVpytor v.1.3.1^104^ with bin size 500. Rare CNVs were defined as those with a population frequency of <5% based on gnomAD v.4.1. CNV annotations were performed manually, consistent with annotations generated by VEP and ANNOVAR, considering only upstream, downstream, 3’ UTR, 5’ UTR, exonic, and intronic regions.

### Analysis of combined effects on gene expression

We assessed for combined effects by first extracting differentially expressed genes (padj<0.05) by DESeq2 in individuals carrying both 16p12.1 deletion and the specific secondary variant compared to all other individuals in the same family. Then, rare variants in those DEGs were extracted and filtered by those which were inherited in the family. Next, two-way ANOVA (using the statsmodels.formula.api and statsmodels.api modules in Python v.3.11.7) was performed using TPM (Transcripts Per Million) of genes annotated from the filtered variants. Genes and variants were kept only when the interaction p value (16p12.1 deletion: secondary hit”, “PR(>F)”) was <0.05 using two-way ANOVA tests. The intersection of the variants with Diffpeak regions was identified using Pybedtools^105^v.0.12.0.

### ATAC-seq sample processing and analysis

Live cell samples were collected and run through a 30 µm filter to obtain a single cell suspension before cryopreservation. Thawed cells were washed and treated with DNAse I (Life Tech, Cat. #EN0521) to remove genomic DNA contamination. Live cell samples were quantified and assessed for viability using a Countess Automated Cell Counter (ThermoFisher Scientific, Waltham, MA, USA). After cell lysis and cytosol removal, nuclei were treated with Tn5 enzyme (Illumina, Cat. #20034197) for 30 minutes at 37°C and purified with Minelute PCR Purification Kit (Qiagen, Cat. #28004) to produce tagmented DNA samples. Tagmented DNA was barcoded with Nextera Index Kit v2 (Illumina, Cat. #FC-131-2001) and amplified via PCR prior to a SPRI Bead cleanup to obtain purified DNA libraries. The sequencing libraries were multiplexed and clustered onto a flow cell on the Illumina NovaSeq X and NovaSeq 6000 according to manufacturer’s instructions by Azenta Life Sciences (South Plainfield, NJ, USA). The samples were sequenced using a 2x150bp Paired End (PE) configuration. Image analysis and base calling were conducted by the NovaSeq Control Software (NCS). Raw sequence data (.bcl files) generated from Illumina NovaSeq was converted into fastq files and de-multiplexed using Illumina bcl2fastq 2.20 software. One mismatch was allowed for index sequence identification. Quality control and processing of fastq sequencing files were performed using the ENCODE ATAC-seq Data Standards and Processing Pipeline (https://github.com/ENCODE-DCC/atac-seq-pipeline). DESeq2 was used to identify differential peaks. The cutoff for adjusted p value (padj) was 0.01. Motif enrichment analysis was performed using the findMotifsGenome.pl from the HOMER^106^ on regions of Diffpeaks.

### ATAC-seq peak Annotation

We annotated ATAC-seq peaks using the union of three approaches: ChIPseeker^107^ the Activity-by-Contact (ABC) model^108^, and nearest gene assignment. ChIPseeker analysis was performed via Galaxy (version 1.28.3), using GENCODE Release 46 (GRCh38.p14) as the reference genome annotation. The ABC model was applied using RNA-seq and ATAC-seq data from HD_01 and HD_02 at the iPSC and NPC stages. Publicly available Hi-C and H3K27ac ChIP-seq datasets were used as additional inputs for contact estimation and enhancer activity scoring^109, 110^. Expressed genes were defined as those with expression >1 TPM and promoter activity quantile >0.4. For each chromosome, enhancer–gene pairs within 5 Mb of a transcription start site (TSS) were scored, and those with an ABC score above 0.022 (default threshold) were retained as predicted regulatory interactions. Peak-to-gene associations were also established by linking the nearest gene to peaks using the closest function from the Pybedtools.

### CRISPRa experiments

iPSCs derived from probands were first infected with lenti dCas9-VP64_Blast and lenti MS2-P65-HSF1_Hygro. Lenti dCAS-VP64_Blast (Addgene plasmid # 61425; http://n2t.net/addgene:61425; RRID: Addgene_61425) and lenti MS2-P65-HSF1_Hygro (Addgene plasmid # 61426; http://n2t.net/addgene:61426; RRID: Addgene_61426)) were both gifts from Feng Zhang. Then, cells were infected with lenti sgRNA (MS2) _Zeo. The EV line was infected with only the backbone. To insert sgRNA, the backbone was digested with BsmBI-v2 (NEB) and ligation was performed using quick ligase (NEB #M2200S) with phosphorylated and annealed sgRNA. Lenti sgRNA (MS2) _Zeo backbone was a gift from Feng Zhang (Addgene plasmid # 61427; http://n2t.net/addgene:61427; RRID: Addgene_61427). sgRNA sequence: *POLR3E*: CACGGCCTGCATGAATGGCG; *MOSMO*: GAGCCGGGAGGACGGAGCTG; *UQCRC2*: ATAAAGAGAGCAGTAGAGCG. All the transductions were performed with 4µg/ml polybrene to improve efficiency. After 24hr incubation, the medium was switched to fresh culture medium and incubated for another 48hr. Selection was then performed at the iPSC stage using 10ng/ml Blasticidin, 250ng/ml Hygromycin, 250ng/ml Zeocin until no further cell death was observed. Selection was repeated using a half dose concentration from day 10 to day 12 of neural rosette formation during the NPC differentiation protocol. Comparing CRISPRa lines to their corresponding EV controls allowed us to identify DEGs resulting from the restoration of specific deletion genes in an isogenic background. These DEGs were subsequently used in downstream functional analyses. Additionally, we defined “reversed genes” as those DEGs from CRISPRa experiments that exhibited changes in the opposite direction compared to DEGs identified in the proband line compared to nondeletion sibling or healthy donor lines.

### PPI network analysis

Protein–protein interaction (PPI) network analysis was performed using the STRING database with default settings (network type: full STRING network; required confidence score: ≥0.400; FDR stringency: medium, 5%). To create the density plot, the active interaction sources were text mining, experiments, databases, co-expression, neighborhood, gene fusion, co-occurrence. Anderson-Darling k-sample test was performed by ad.test function in kSamples v1.2-10 package in R. To visualize the proteins involved in signal transduction based on Reactome data, active interaction sources were experiments, databases and co-expression. Average node degree, PPI enrichment p-value and functional enrichment were obtained from the STRING database.

### Enrichment analysis

We performed over-representation analysis (ORA) and gene set enrichment analysis (GSEA) using clusterProfilter v.4.12.6 package in R, following the described guidelines^111^. For ORA, we used enrichGO, entichKEGG, and enrichPathway functions. For GSEA, analysis was performed against curated gene sets obtained using the msigdbr v.10.0.1 package in R. Pathway terms belonging to Reactome, WP, and KEGG databases were extracted in an unbiased manner. For generation of the waterfall plots in Figure 3G and Supplementary Figure 6D, the top 30 pathways were unbiasedly selected from each database at the iPSC, NPC, iMN, and MN stages. These pathways were then pooled, and the top 100 were selected based on q-value. If the number of significantly enriched terms was fewer than 30 at each stage or fewer than 100 in total after pooling, all enriched terms were included for generating waterfall plots. Enrichment analysis of DEGs in published gene lists was performed using the GeneOverlap v.1.36.0^112^ package in R. Enrichment analysis of DEGs in DisGeNET was performed using enrichR v.3.4^113^ package in R.

### Construction of TF regulatory networks for JUN and FOXG1

We used the ChEA3 (https://maayanlab.cloud/chea3/) web application to construct TF–TF co-regulatory networks using the top 15 TFs whose binding motifs were significantly enriched in Diffpeak regions from deletion lines versus HD lines comparisons at the NPC stage. In comparisons where fewer than 15 motifs were enriched, only motifs with p-value less than 0.01 were included. FOXG1 and JUN ranked highest among the enriched transcription factors based on the Integrated Scaled Rank. TF-TF co-regulatory networks were constructed by ChEA3 using the top results, and edges between TFs defined by the evidence from libraries which supports the interactions. To identify regulatory network genes for each TF, we input DEGs from each comparison into ChEA3 and used the “Overlapping Genes” column. The top integrated rank across ChEA3 libraries was used in our analysis.

### Imaging and quantification

Revolve (Discover Echo) was used to detect Click-iT signals. LSM 800 upright and LSM 800 inverted (ZEISS) were used to detect fluorescence signals for immunofluorescence assays. Images were analyzed using Fiji (ImageJ).

### Statistics and reproducibility

Investigators were blinded to experimental condition when quantifying fluorescence intensity. Comparisons between deletion carrier and noncarrier lines within the same family were used when possible to control for the independent effects of shared secondary variants and to specifically analyze the joint effects caused by both the 16p12.1 deletion and secondary variants. Comparisons with healthy donor lines provided a consistent baseline across families and served as appropriate controls for cellular assays and functional analysis, as many noncarrier lines also exhibited clinical features due to the underlying genetic liability. The same bioinformatics pipelines were applied across all conditions for each assay. Statistical analyses were conducted using R v4.3.1 and Python v3.11.7. Details of the statistical tests and number of replicates are provided in the corresponding figure legends and Supplementary Tables.

## Data Availability

Data will be deposited to dbGaP. The data will consist of WGS, RNA-seq, and ATAC-seq data. All processed data are presented as supplementary tables.

## Acknowledgments

The authors thank Dr. Yingwei Mao for valuable advice on iPSC culture and Dr. Melissa Rolls for providing imaging resources. We are grateful to the National Institute of Neurological Disorders and Stroke (NINDS) for supplying iPSC lines derived from healthy donors. The authors thank Dr. Matthew Jensen, Dr. Francisca Canzar and Johnathan Ray for their assistance and insights in data analysis. The following funding supported this study: NIH R01GM121907 and R21NS122398 (to S.G.)

## Author contributions

J.S. and S.G. conceived of and designed the study. S.G. supervised the experiments and analyses. J.S., S.N. and M.D. maintained iPSC culture, performed the neural conversion and extracted RNA. J.S., M.D. and A.P. generated the CRISPR-edited lines. J.S. and S.N. performed immunostaining, imaging and image quantification. J.S., and V.H.B. performed assays on cell proliferation and apoptosis. J.S. designed CRISPRa experiments and J.S., V.H.B. performed CRISPRa experiments. J.S., D.B., C.S., and B.G. performed analyses on ATAC-seq, RNA-seq and WGS. C.S. summarized the clinical phenotype information. D.A. and K.P. recruited 16p12.1 deletion families. P.L. reprogrammed iPSCs from PBMCs derived from recruited patients. J.S., S.N. and S.G. wrote the paper.

## Data and code availability

Whole genome sequencing, RNA-seq and ATAC-seq generated in this study are available at NCBI dbGaP phs002403.v1.p1. All code generated for this project, including pipelines for running bioinformatic software and custom analysis scripts, are available at https://github.com/Jiawan1023/iPSC_integrated_framework. Detailed information of statistical analyses is available in Supplementary Tables.

**Supplementary Table 1:**

Lists of DEGs and altered isoforms with secondary variants in the isogenic setting; Detailed results of Fisher’s exact tests in Figure 1C and volcano plots in Supplementary Figure 2A.

**Supplementary Table 2:**

Lists of Diffpeaks with secondary variants in the isogenic setting; Detailed results of Fisher’s exact tests in Figure 1D and volcano plots in Supplementary Figure 2B.

**Supplementary Table 3:**

Lists of TPM values of combined effects on gene expression across iPSC, NPC, iMN and MN stages in the familial setting; Detailed results for Figure 1C.

**Supplementary Table 4:**

Lists of DEGs, reversed genes, GSEA results and ORA results in CRISPRa experiments.

**Supplementary Table 5:**

Lists of DEGs with secondary variants, and DEGs with secondary variants involved in signal transductions for Figure 5.

**Supplementary Table 6:**

Lists of enriched motifs at the NPC stage, DEGs and Diffpeaks for Supplementary Figure 10B and ChEA3 result.

**Supplementary Table 7:**

Lists of detailed results for WGCNA.

**Supplementary Table 8:**

Lists of DESeq2 results and GSEA results for each comparison, enrichment results using published datasets and DisGeNET.

**Supplementary Table 9:**

Summary of all statistical tests.

---

**Supplementary Figure 1.**
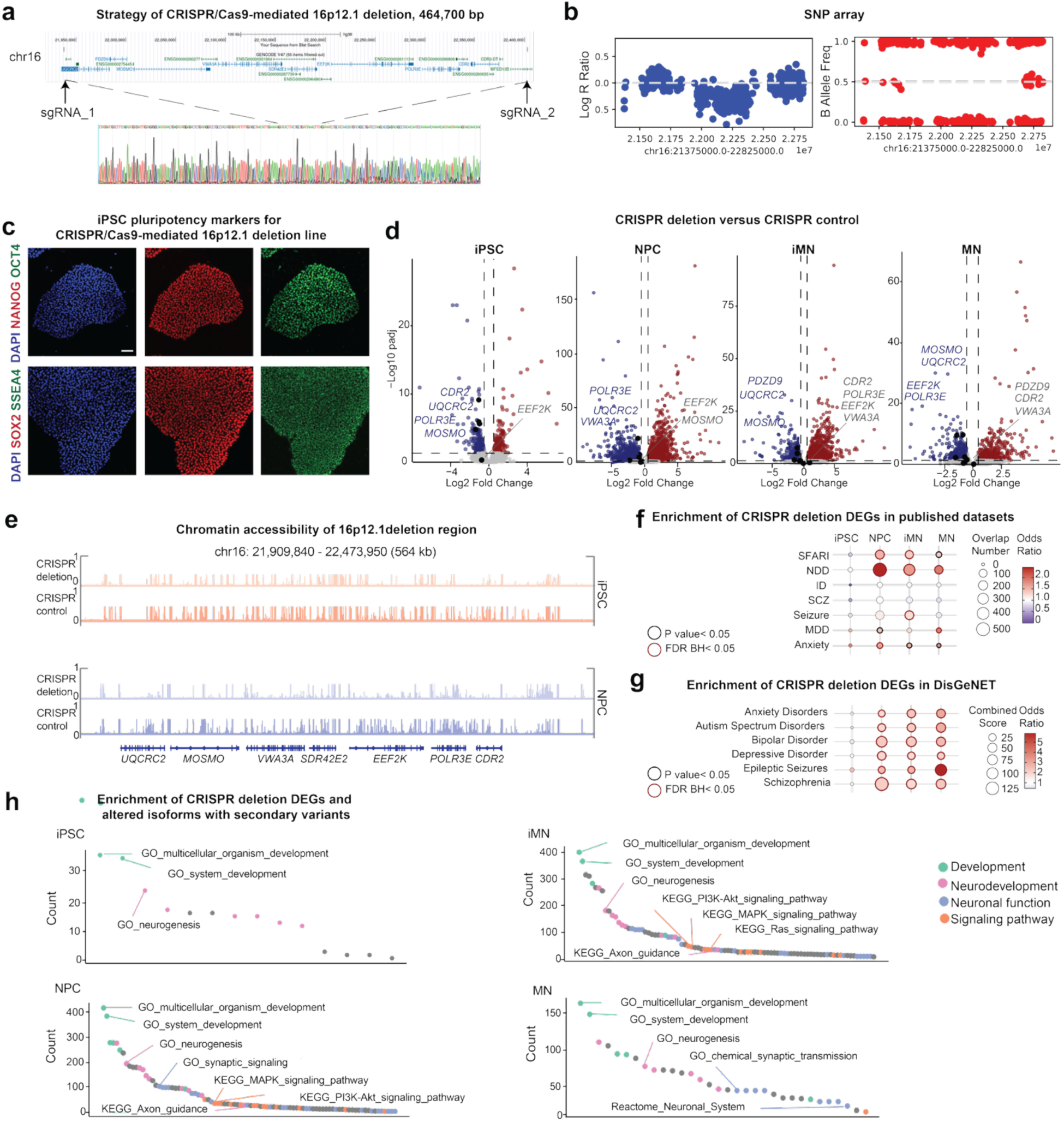
Generation of CRISPR/Cas9-mediated 16p12.1 deletion. **(a)** CRISPR/Cas9 strategy for generating 16p12.1 deletion. Arrows demonstrate the locations of the designed sgRNAs. The engineered 465-kbp deletion was validated by Sanger sequencing. **(b)** Confirmation of the deletion by SNP array. **(c)** Representative images of the CRISPR edited line stained for pluripotency markers (SOX2, SSEA4, NANOG and OCT4). Scale bars, 70 μm. **(d)** Volcano plots show the differential expression of 16p12.1 deletion genes (*UQCRC2*, *MOSMO*, *VWA3A*, *EEF2K*, *POLR3E* and *CDR2*) in the isogenic setting across neuronal differentiation stages (iPSC, NPC, IMN and MN). Dashed lines represent the threshold (log_2_|FC|≥0.5, padj<0.05). Red, upregulated genes; blue, downregulated genes; grey, non-significant genes. Deletion genes that are not labeled demonstrate extremely low expression levels and were not detected. **(e)** IGV snapshots show the ATAC-seq peaks of chromatin accessibility within the 16p12.1 deletion region from the CRISPR-edited line. **(f)** Enrichment analysis of DEGs (log_2_|FC|≥1, padj <0.05) obtained from RNA-seq of the CRISPR-edited line compared to isogenic controls queried within published datasets. Gene lists from selected papers are provided in **Supplementary Table 8**. **(g)** Enrichment analysis of DEGs (log_2_|FC|≥1, padj <0.05) obtained from RNA-seq of the CRISPR-edited line compared to isogenic controls within DisGeNET. For **(f)** and **(g)**, Fisher’s exact test, bold black circles indicate p<0.05, bold red circles indicate Benjamini-Hochberg FDR<0.05. **(h)** Waterfall plot shows enrichment of DEGs overlapping secondary variants across the 4 neuronal differentiation stages. Top 30 terms (q<0.05) were unbiasedly selected from GO (Biological Process, Gene Ontology), KEGG (Kyoto Encyclopedia of Genes and Genomes) and Reactome databases. RNA-seq and downstream analyses: n=3 independent experiments; ATAC-seq and downstream analyses: n=3 independent experiments.

**Supplementary Figure 2.**
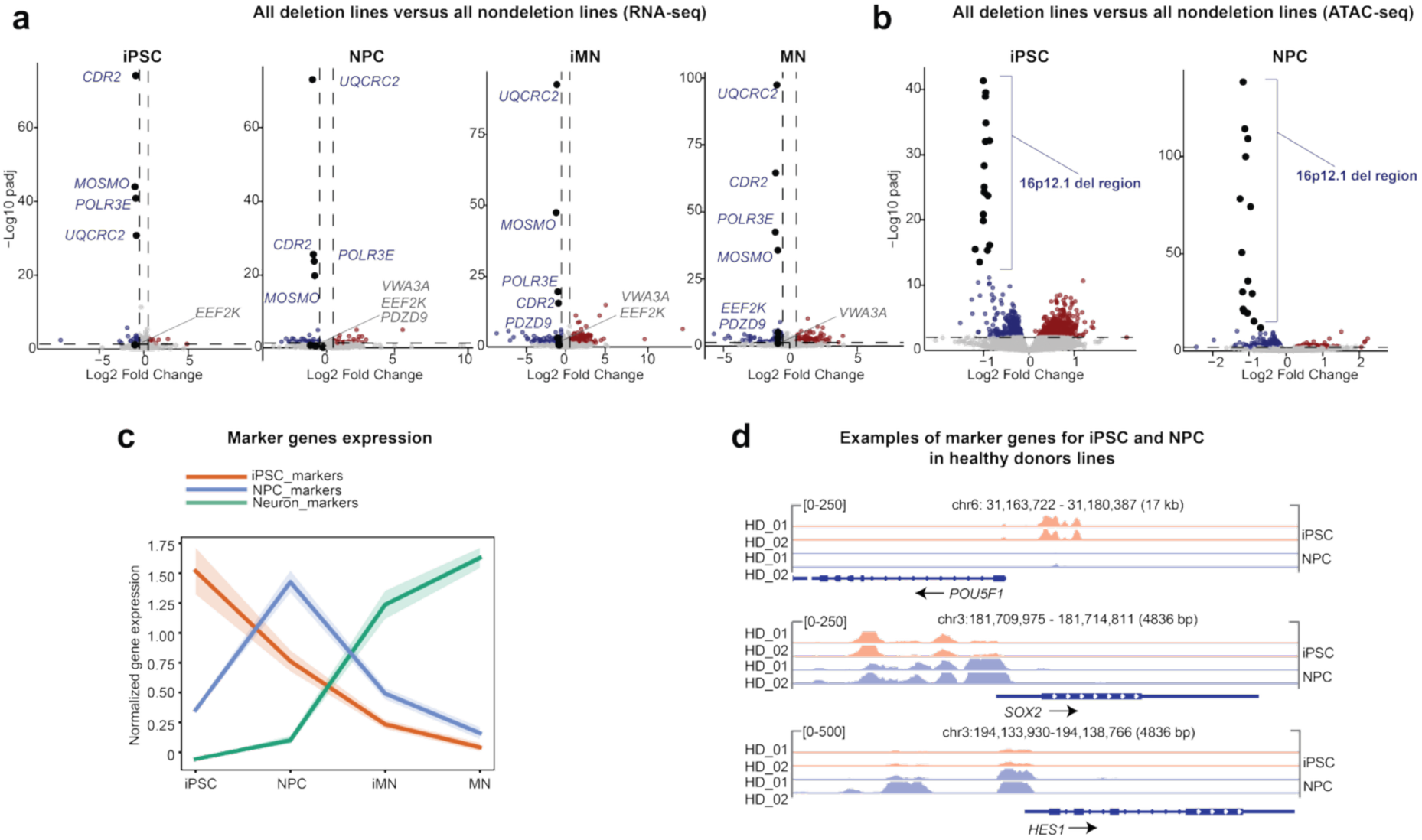
Validation of cell identities across stages of neuronal differentiation. **(a)** Volcano plots show the differential expression of 16p12.1 deletion genes (*UQCRC2*, *MOSMO*, *VWA3A*, *EEF2K*, *POLR3E* and *CDR2*) comparing all the deletion lines to nondeletion lines across neuronal differentiation stages (iPSC, NPC, IMN and MN). Dashed lines represent the statistical threshold for significance (log_2_|FC|≥0.5, padj<0.05). Red, upregulated genes; blue, downregulated genes; grey, non-significant genes. Deletion genes that are not labeled demonstrate extremely low expression levels and were not detected. **(b)** Volcano plots show the differential peaks in 16p12.1 deletion when comparing all the deletion lines to nondeletion lines at iPSC and NPC stages. Dashed lines represent the threshold (padj<0.01). Red, upregulated peaks; blue, downregulated peaks; grey, non-significant peaks. Black dots demonstrate differential peaks within 16p12.1 regions. A list of all ATAC-seq peak regions is provided in **Supplementary Table 2**. **(c)** Line plot shows the expression of select marker genes with shading representing 95% confidence interval of the mean across all the lines and differentiation stages. Y-axis represents z-scores of TPM. iPSC markers: SOX2, POU5F1, and FUT4; NPC markers: SOX2, HES1, NES, and EMX2; neuron markers: MAP2, DLG4, NCAM1, and RBFOX3. **(d)** IGV snapshots show the ATAC-seq peaks of chromatin accessibility from HD_01 and HD_02 in the 16p12.1 deletion regions. For RNA-seq, ATAC-seq and downstream analyses: n=3 independent experiments, except for RNA-seq and downstream analyses where n=2 for iMNs and MNs of P1C_077, and MNs of HD_01.

**Supplementary Figure 3.**
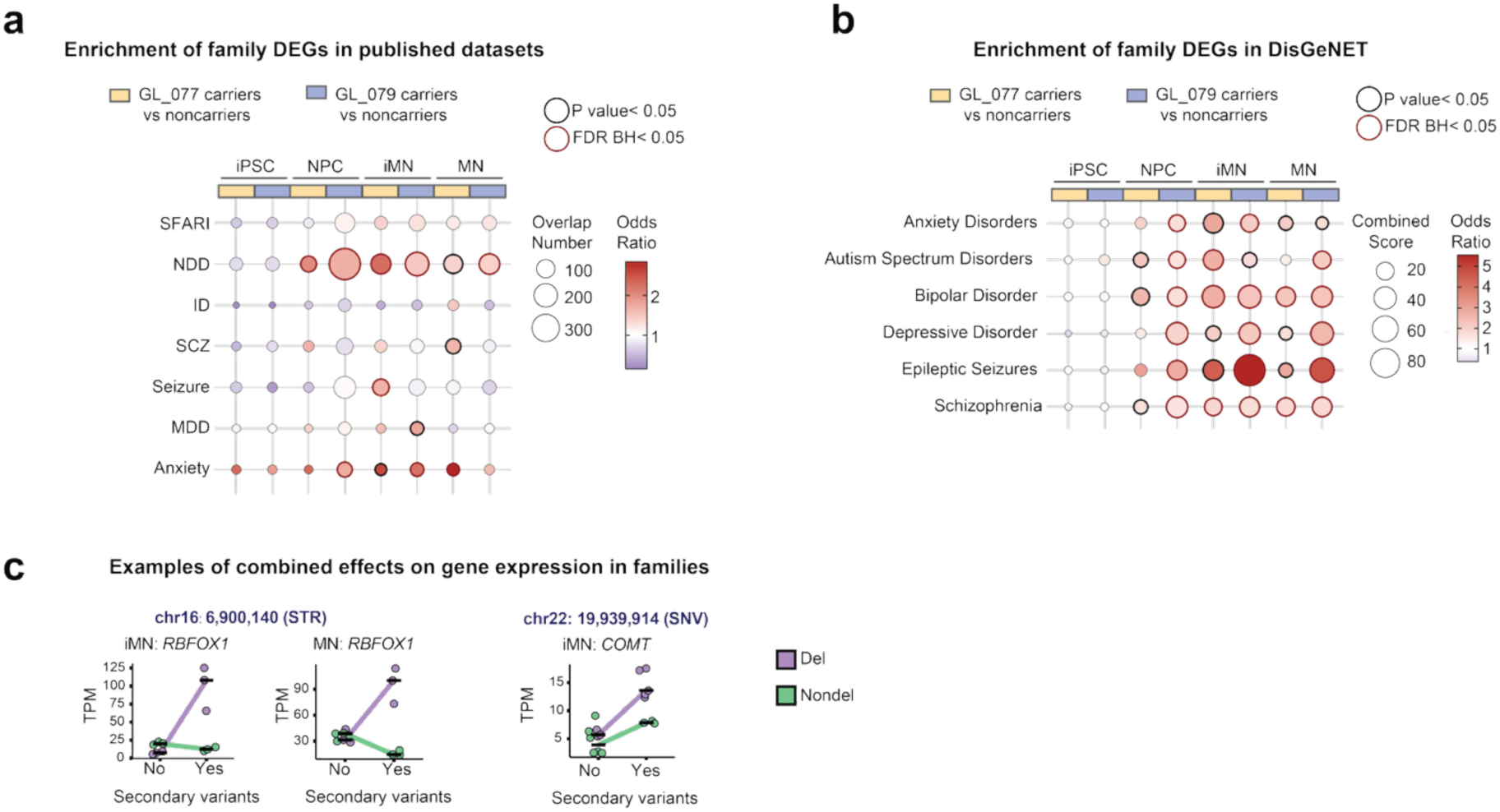
Family-specific effects on gene expression. **(a)** Enrichment of DEGs (log_2_|FC|≥1, padj <0.05) from deletion versus nondeletion within-family comparisons in published datasets. A complete list of genes from select papers is provided in **Supplementary Table 8**. **(b)** Enrichment analysis of DEGs (log_2_|FC|≥1, padj <0.05) from deletion versus nondeletion within-family comparisons in DisGeNET. For **(a)** and **(b)**, Fisher’s exact test, bold black circles indicate p<0.05, bold red circles indicate Benjamini-Hochberg FDR<0.05. GL_007 carriers are MC_077 and P1C_077, noncarriers are FNC_007 and S2NC_077, and GL_079 carriers are MC_079 and P2C_079, noncarriers are FNC_079 and S2NC_079. **(c)** In the same family, examples of genes and isoforms overlapping with secondary variants showing altered (TPM and mean values) gene expression at iMN and MN stages (**Supplementary Table 3**). *indicates the carriers of secondary variants. SNV, single nucleotide variant; STR, short tandem repeat; Del, 16p12. 1 deletion lines; Nondel, nondeletion lines. Two-way ANOVA tests for all the examples using TPM are interaction p<0.05. For RNA-seq and downstream analyses: n=3 independent experiments, except for iMNs and MNs of P1C_077, where n=2.

**Supplementary Figure 4.**
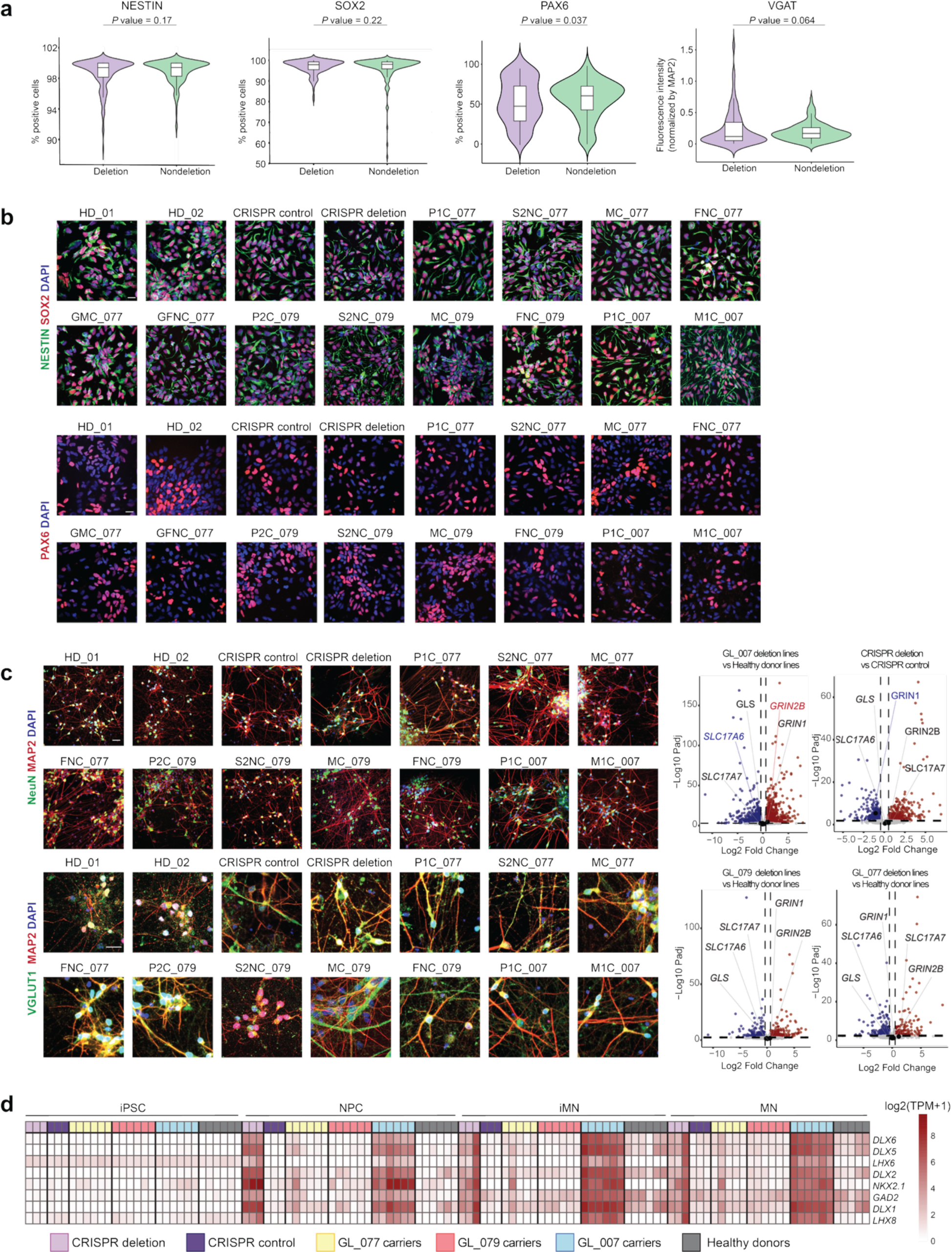
Characterization of NPCs and neuronal lineage markers. **(a)** Violin plots show the percentage of NPCs that were positive for NESTIN, SOX2, PAX6 and quantification of VGAT staining in neurons from all deletion lines versus all nondeletion lines. The embedded box plot indicates the interquartile range (IQR) and median (black horizontal line). p-values calculated using two-tailed Welch’s t-test. **(b)** Representative images of NPCs stained for NESTIN, SOX2, and PAX6. **(c)** Representative images (left) of MNs stained for NeuN and MAP2 (mature neuron markers) and VGLUT (excitatory neuron marker), and volcano plots (right) showing the differential expression of five marker genes for glutamatergic neurons between deletion lines in each family and healthy donor lines, as well as the CRISPR deletion compared to the CRISPR control. Red, upregulated genes; blue, downregulated genes; grey, non-significant genes. **(d)** Heatmap shows the expression levels of inhibitory neuron-related genes across neuronal differentiation stages. Note that the differences in gene expression between the CRISPR-edited line and controls become more subtle as neuronal differentiation progresses (from iPSC to MN). Color scale represents log₂(TPM+1). RNA-seq and downstream analyses: n=3 independent experiments, except for MNs of HD_01, where n=2, and iM and MNs of P1C077, where n=2. Scale bars for **(b)** and **(d)**, 20 μm.

**Supplementary Figure 5.**
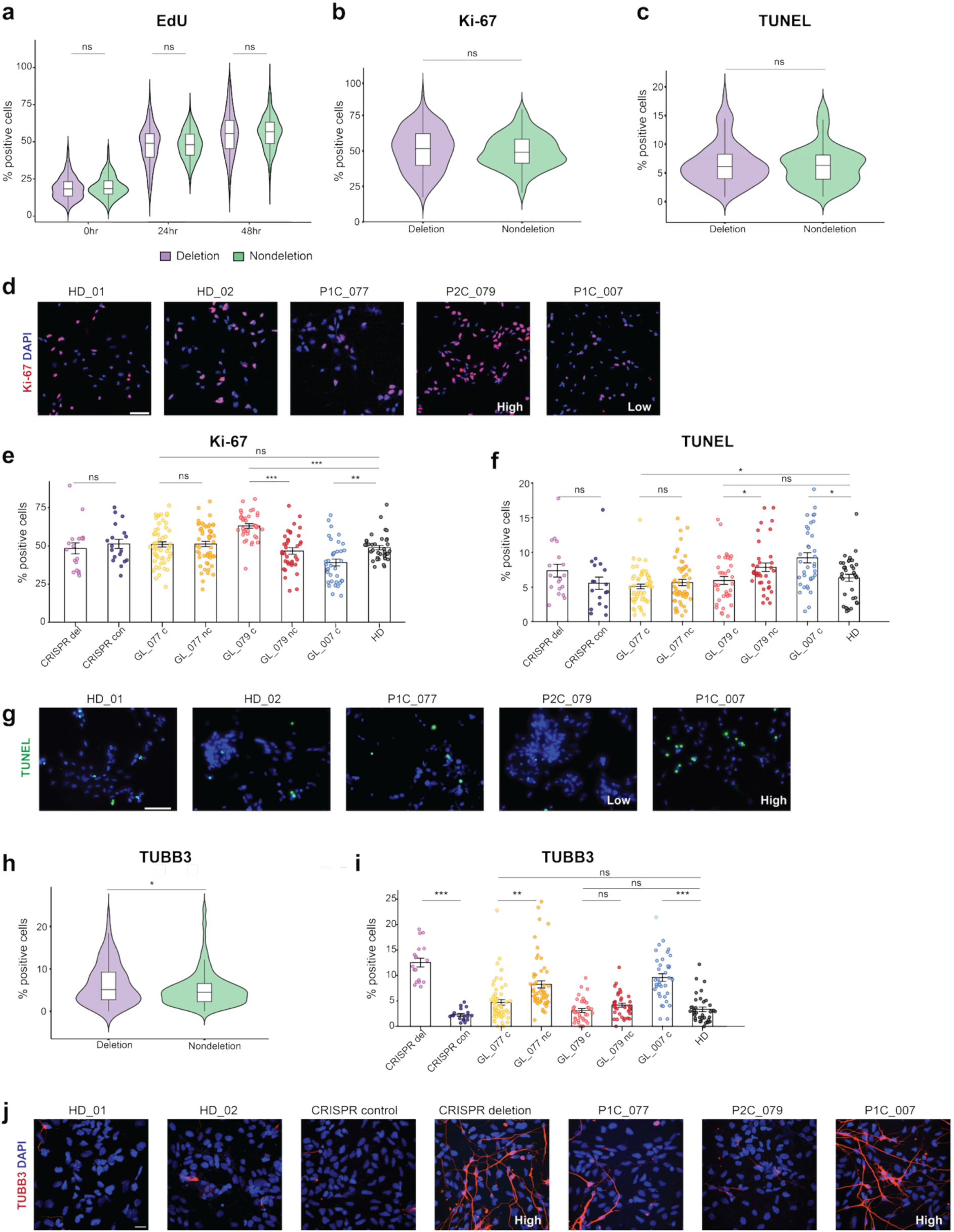
Altered proliferation, apoptosis, and neuronal maturation at the NPC stage. **(a)** Violin plot shows the percentage of EdU-positive cells at the NPC stage in all deletion lines versus all nondeletion lines across three time points. The embedded box plot indicates the interquartile range (IQR) and median (black horizontal line). **(b)** Violin plot shows the percentage of Ki-67 positive cells at the NPC stage in all the deletion lines versus all the nondeletion lines. The embedded box plot indicates the IQR and median (black horizontal line). **(c)** Violin plot shows the percentage of TUNEL-positive cells at the NPC stage in all the deletion lines versus all the nondeletion lines. The embedded box plot indicates the interquartile range (IQR) and median (black horizontal line). **(d)** Representative images of NPCs stained for Ki-67. Scale bars, 70 μm. **(e)** Bar plot (with mean values) shows the percentage of Ki-67-positive cells in different samples. **(f)** Bar plot shows the percentage of TUNEL-positive cells in different samples. **(g)** Representative images of TUNEL-labeled NPCs. Scale bars, 70 μm. **(h)** Violin plot shows the percentage of TUBB3-positive cells at the NPC stage in all the deletion lines versus all the nondeletion lines. The embedded box plot indicates the interquartile range (IQR) and median (black horizontal line). **(i)** Bar plot shows the percentage of TUBB3-positive cells in different samples. **(j)** Representative images of TUBB3-positive NPCs. Scale bars, 20 μm. CRISPR_del, CRISPR deletion; CRISPR_con, CRISPR control; GL_077_c, 16p12.1 deletion lines in GL_077 (GMC_077, MC_077, P1C_077); GL_077_nc, nondeletion lines in GL_077 (GFNC_077, FNC_077, S2NC_077); GL_079_c, deletion lines in GL_077 (MC_079, P2C_079); GL_079_nc (FNC_079, S2NC_079); GL_007_c (M1C_007, P1C_007); HD, healthy donor lines (HD_01, HD_02). Two-tailed Welch’s t-test, *p<0.05; **p<0.01; ***p<0.001; ****p<0.0001, for **(a)**, **(b)**, **(c)**, **(e)**, **(f)**, **(h)**, **(i)**. n=3 independent experiments for each line. 6 images per experiment were quantified.

**Supplementary Figure 6.**
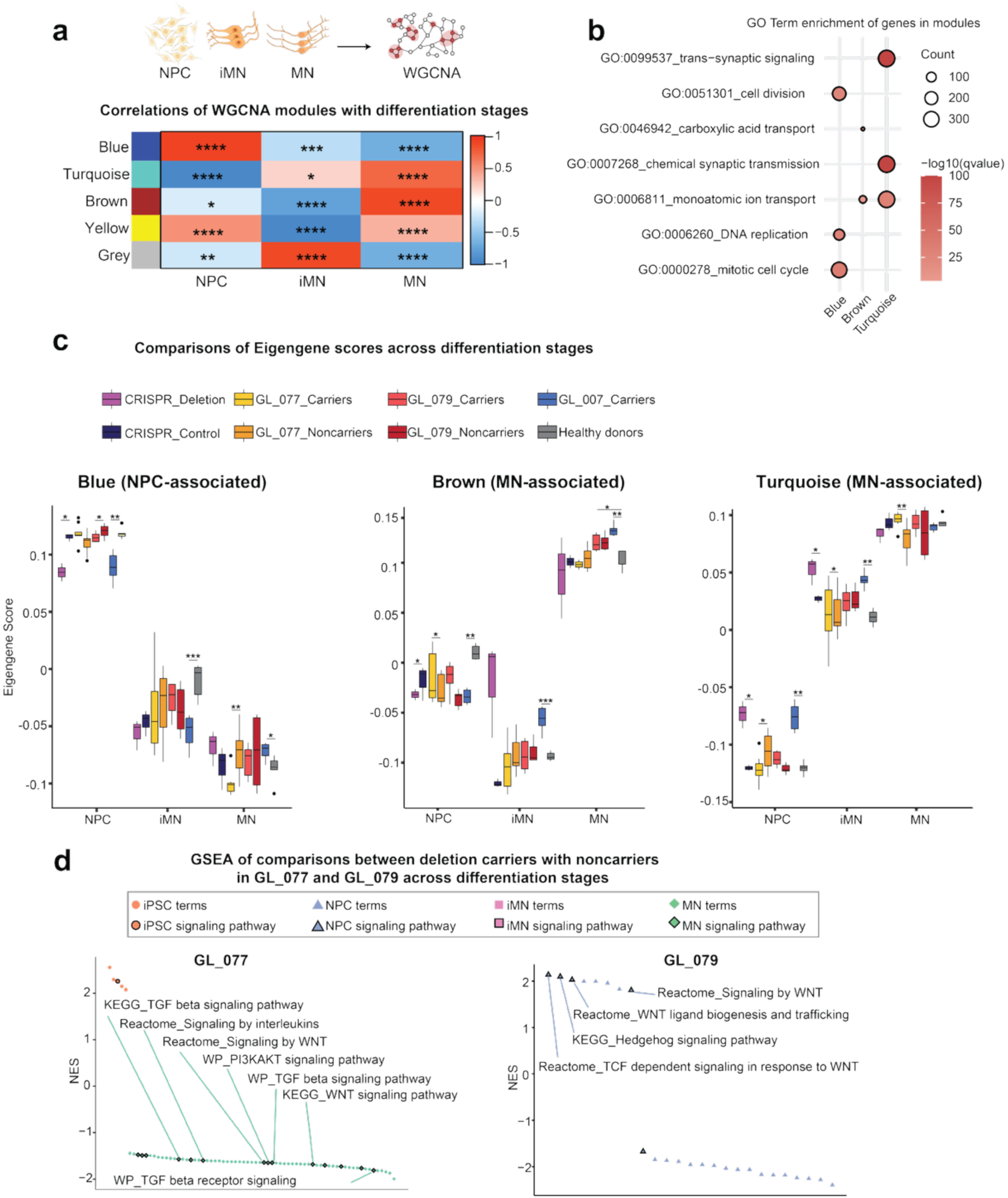
Transcriptome dynamics during neuronal differentiation. **(a)** Heatmap shows Pearson’s correlation coefficients between WGCNA modules and differentiation stages. Student asymptotic p-value for correlation for statistics of Pearson correlation. *p<0.05; **p<0.01; ***p<0.001; ****p<0.0001. Color bar represents direction and strength of correlation. **(b)** GO term enrichment of genes in each module. **(c)** Box plots show the eigengene scores across the differentiation stages. Two-tailed Welch’s t-test. *p<0.05; **p<0.01; ***p<0.001; ****p<0.0001. Each box shows the IQR, with a horizontal line indicating the median. **(d)** Waterfall plot of total unbiasedly selected top 100 pathways enriched for the DEGs identified across differentiation stages comparing deletion to nondeletion lines in GL_077 and GL_079 by GSEA (q<0.05) annotated in Reactome, KEGG and WP (WikiPathway) databases. The bold border indicates terms that belong to signaling pathways. NES, normalized enrichment score; GL_077, GMC_077, MC_077, and P1C_077 versus GFNC_077, FNC_077, and S2NC_077; GL_079, MC_079, and P2C_079 versus FNC_079 and S2NC_079. n=3 independent experiments, except for RNA-seq and downstream analyses where n=2 for iMNs and MNs of P1C_077, and MNs of HD_01.

**Supplementary Figure 7.**
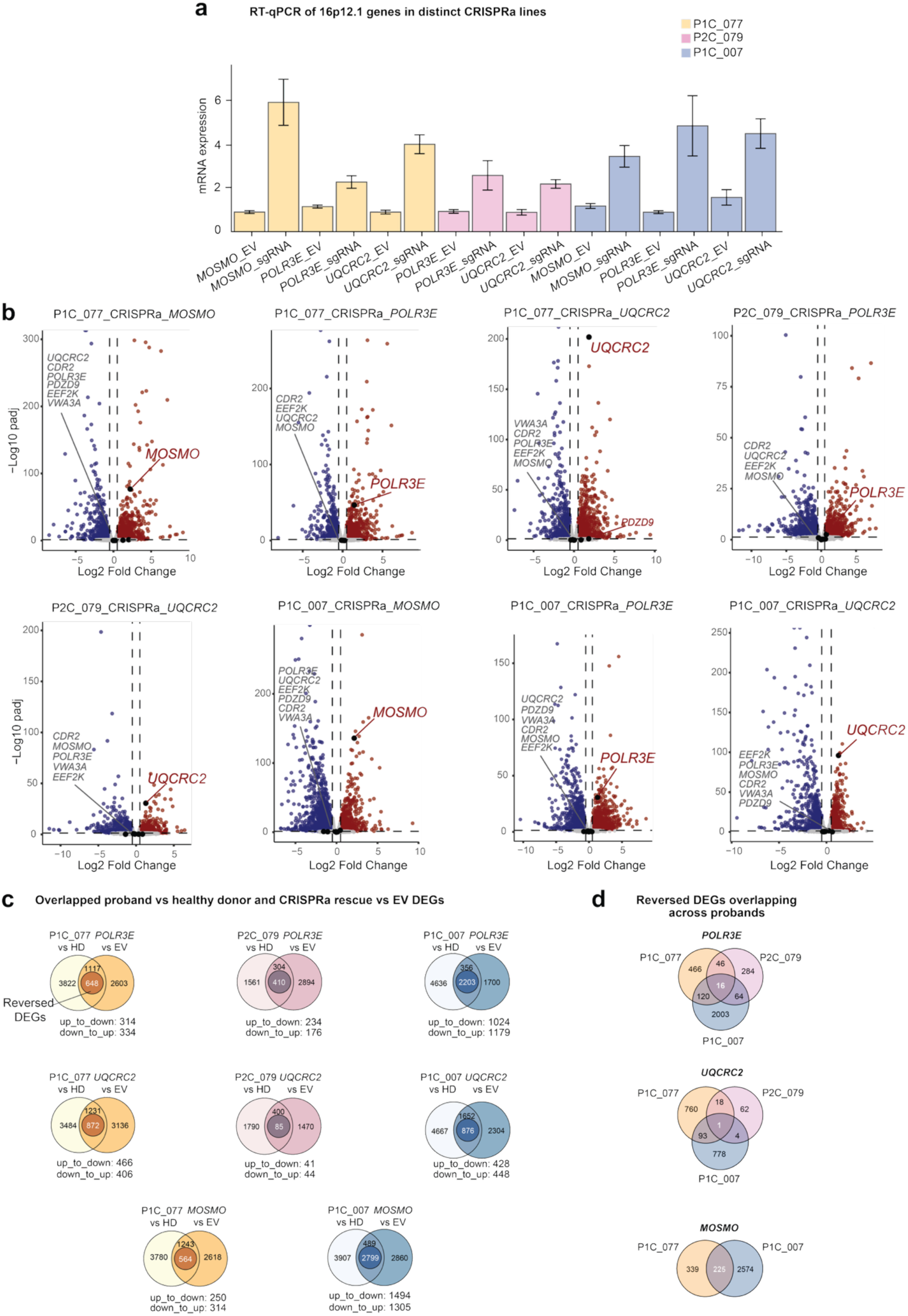
Transcriptomic alterations induced by CRISPRa-mediated activation of individual 16p12.1 genes. **(a)** Bar plot shows relative mRNA expression levels of *MOSMO*, *POLR3E*, and *UQCRC2* across CRISPRa lines derived from the three probands (P1C_077, P2C_079, and P1C_007), measured by RT-qPCR. Each bar represents the mean±standard error of three independent experiments. **(b)** Volcano plots show the differential expression of 16p12.1 deletion genes (*UQCRC2*, *MOSMO*, *VWA3A*, *EEF2K*, *POLR3E*, and *CDR2*) comparing CRISPRa of individual 16p12.1 genes to EV lines. Dashed lines represent the statistical significance threshold (log_2_|FC|≥0.5, padj<0.05). Red, upregulated genes; blue, downregulated genes; grey, non-significant genes. Deletion genes that are not labeled demonstrate extremely low expression levels and were not detected. **(c)** Venn diagrams show the overlaps (reversed DEGs) of DEGs (probands with the deletion versus HDs) and DEGs (padj<0.05) from CRISPRa of individual genes versus EV in the proband lines. Gene lists are provided in **Supplementary Table 4**. **(d)** Venn diagrams show the patterns of all DEGs that were reversed by CRISPR activation of each 16p12.1 gene across probands. Gene lists are provided in **Supplementary Table 4**. Up_to_down, upregulated in deletion lines but downregulated by CRISPRa; down_to_up, downregulated in deletion lines but upregulated by CRISPRa; HD, healthy donors (HD_01 and HD_02); EV, empty sgRNA-MS2 vector.

**Supplementary Figure 8.**
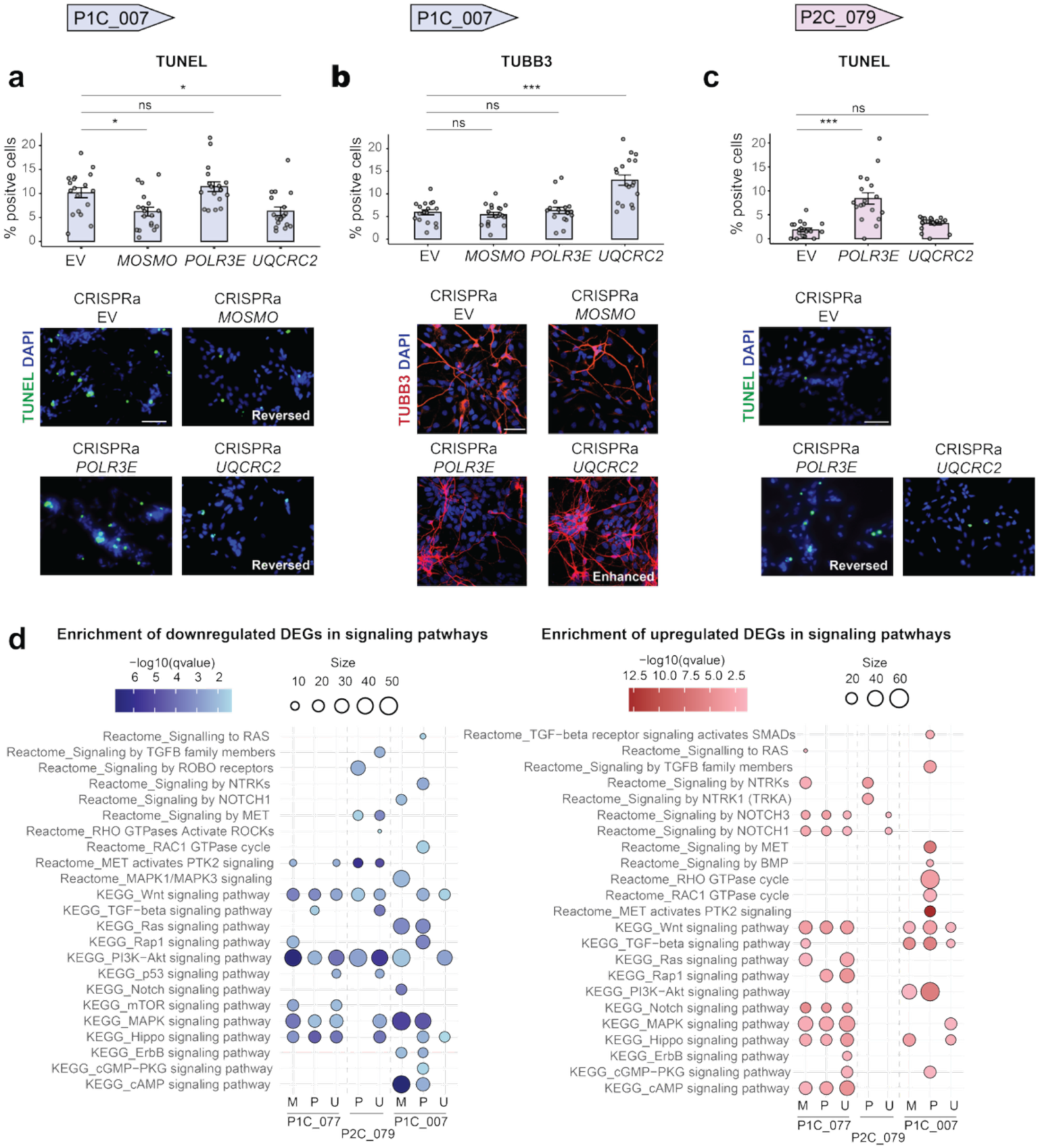
Alterations in cellular phenotypes following CRISPR activation of 16p12.1 genes. **(a)** Quantification and representative images of TUNEL-labeled NPCs in CRISPR activated lines from P1C_007. Scale bars, 70 μm. **(b)** Quantification and representative images of NPCs stained for TUBB3 in CRISPR activated lines from P1C_007. Scale bars, 20 μm. **(c)** Quantification and representative images of TUNEL-labeled NPCs in CRISPR activated lines from P2C_079. Scale bars, 70 μm. **(d)** Plots show enrichment of downregulated (left, blue) and upregulated (right, red) DEGs within signaling pathways (log_2_|FC|≥0.5, padj <0.05) obtained from RNA-seq of CRISPR activated versus EV lines in probands (M: *MOSMO*; P: *POLR3E*, and U: *UQCRC2*). The size of the circles represents gene counts within each pathway, and the color intensity corresponds to significance of enrichment. Data are mean±s.e.m and one-way ANOVA followed by Dunnett’s post hoc test, *p<0.05; **p<0.01; ***p<0.001; ****p<0.0001, for **(a)**, **(b)**, and **(c)**. EV, empty sgRNA-MS2 vector.

**Supplementary Figure 9.**
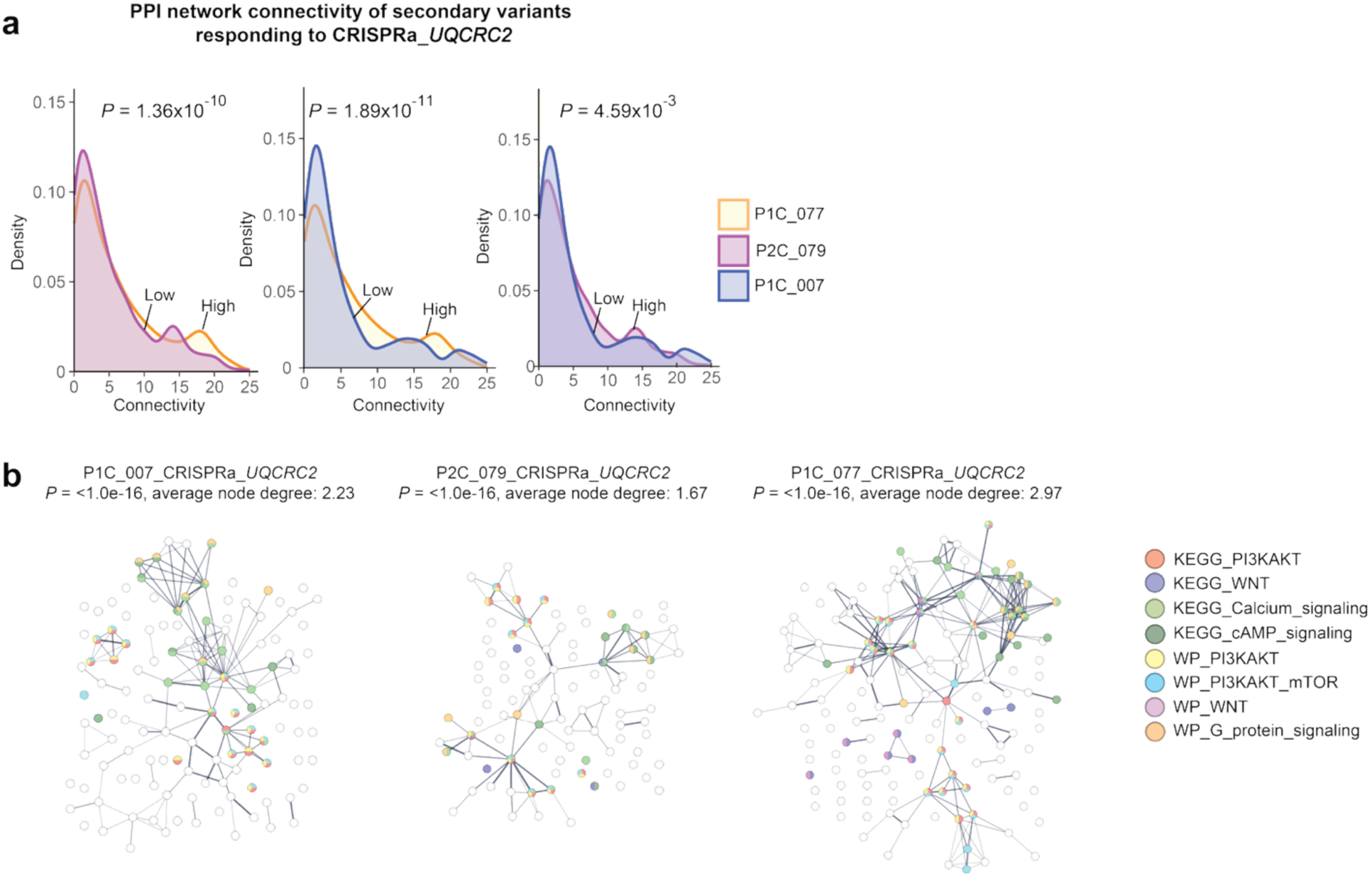
Connectivity of secondary variants in response to CRISPR activation of *UQCRC2* within protein-protein interaction networks. **(a)** Density plots show the connectivity (node degree) of DEGs that overlap with secondary variants (log_2_|FC|≥0.5, padj<0.05) and are reversed with CRISPR activation of *UQCRC2* in three probands, as measured using the STRING database. Anderson-Darling k-sample tests were used to calculate p-values. **(b)** PPI network of DEGs (log₂|FC|≥0.5, padj<0.05) involved in signaling pathways (from Reactome database) that overlap with secondary variants and were reversed by CRISPRa in the probands. For all networks, p<10^-16^. Protein nodes involved in specific pathways were differently colored using the STRING database. Average node degrees and PPI enrichment p values were also calculated using the STRING database.

**Supplementary Figure 10.**
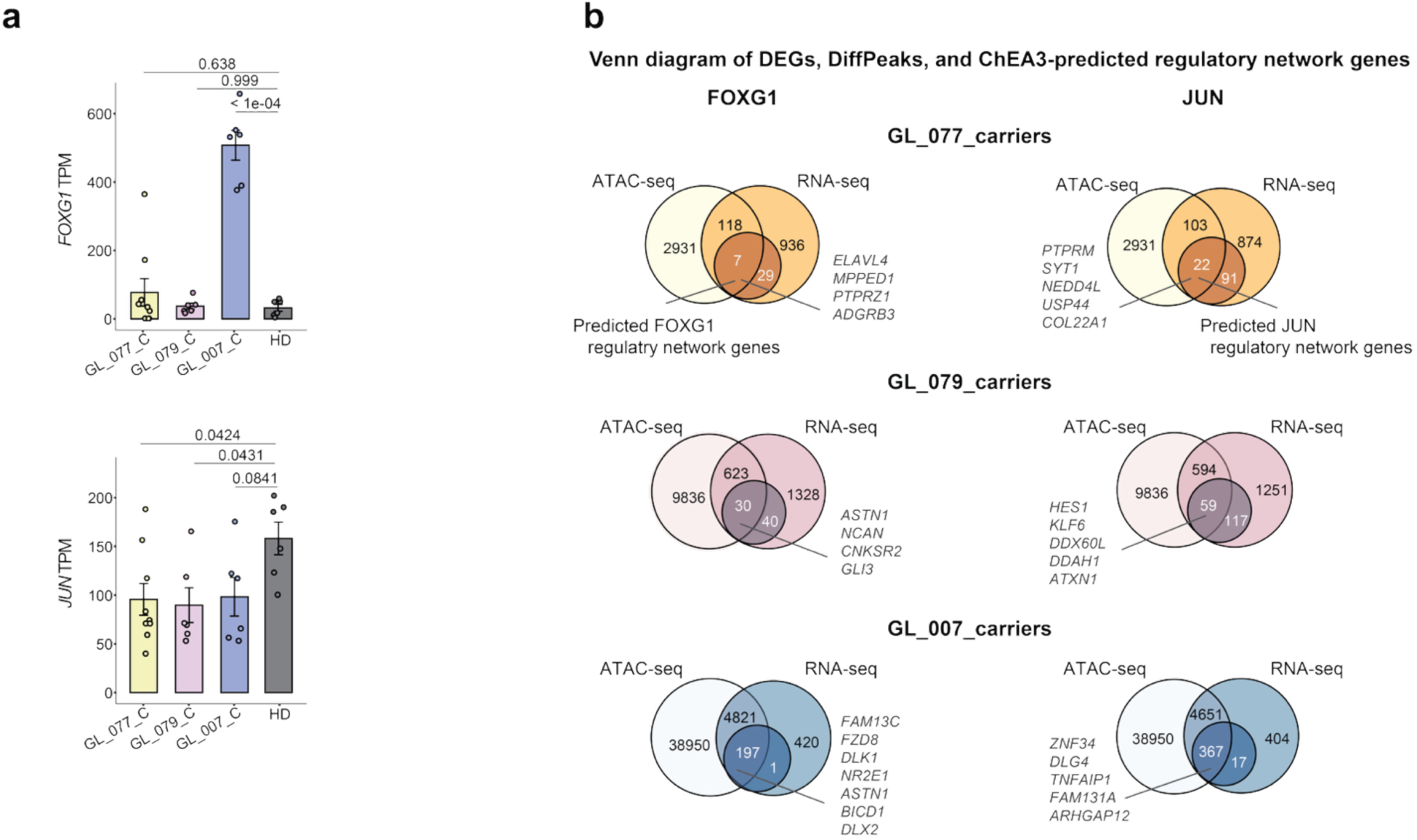
Expression changes of *FOXG1*, *JUN* and their regulatory network genes in NPCs. **(a)** Bar plots with mean values show the TPM of gene expression for *FOXG1* and *JUN* across the proband lines (bottom panel). One-way ANOVA followed by Dunnett’s post hoc test. **(b)** Venn diagrams show the overlap of genes from DEGs (log_2_|FC|≥0.5, padj<0.05), ATAC-seq (padj<0.01), and genes within regulatory networks of FOXG1 and JUN as predicted by ChEA3 based on DEGs. Select example genes are also labeled. GL_077 carriers (GMC_077, MC_077 and P1C_077); GL_079 carriers (MC_079 and P2C_079); GL_007 carriers (M1C_07 and P1C_07); healthy donor (HD_01 and HD_02). n=3 independent experiments.

